# Serum metabolomics improve risk stratification for incident heart failure

**DOI:** 10.1101/2023.08.21.23294202

**Authors:** Rafael R. Oexner, Hyunchan Ahn, Konstantinos Theofilatos, Ravi A. Shah, Robin Schmitt, Philip Chowienczyk, Anna Zoccarato, Ajay M. Shah

## Abstract

**Background and Aims:** Prediction and early detection of heart failure (HF) is crucial to mitigate its impact on quality of life, survival, and healthcare expenditure. In this study, we explored the predictive value of serum metabolomics (168 metabolites detected by proton nuclear magnetic resonance (^1^H-NMR) spectroscopy) for incident HF.

**Methods:** We leveraged data of 68,311 individuals and > 0.8 million person-years of follow-up from the UK Biobank (UKB) cohort to assess individual metabolite associations and to train models to predict HF risk in individuals not previously considered at risk. Specifically, we (I) fitted per-metabolite COX proportional hazards (COX-PH) models to assess individual metabolite associations and (II) trained and internally validated elastic net (EN) models to predict incident HF using the serum metabolome. We benchmarked discriminative capacities against a comprehensive, well-validated clinical risk score (Pooled Cohort Equations to Prevent HF, PCP-HF^1^).

**Results:** During median follow-up of ≈ 12.3 years, several metabolites showed independent association with incident HF (90/168 adjusting for age and sex, 48/168 adjusting for PCP-HF; false discovery rate (FDR)-controlled P < 0.01). Performance-optimized risk models effectively retained key predictors representing highly correlated clusters (≈ 80 % feature reduction). The addition of metabolomics to PCP-HF improved predictive performance (Harrel’s C: 0.768 vs. 0.755.; continuous net reclassification improvement (NRI) = 0.287; relative integrated discrimination improvement (IDI): 17.47 %). Simplified models including age, sex and metabolomics performed almost as well as PCP-HF (Harrel’s C: 0.745 vs. 0.755, continuous NRI: 0.097, relative IDI: 13.445 %). Risk and survival stratification was improved by the integration of metabolomics.

**Conclusions:** The assessment of serum metabolomics improves incident HF risk prediction. Scores based simply on age, sex and metabolomics exhibit similar predictive power to clinically-based models, potentially offering a cost- and time-effective, standardizable, and scalable single-domain alternative to more complex clinical scores.

## Introduction

Heart failure (HF) is a major global health problem associated with high morbidity and mortality. Rising life expectancy, absolute population growth, detrimental lifestyle changes and improved survival after diagnosis all contribute to an increasing prevalence of HF^2–5^. The clinical management of HF is complex, encompassing multimodal therapeutic strategies including improved surveillance and patient education, pharmacotherapy, device implantation and invasive/surgical approaches. Whilst valuable for affected individuals, these therapeutic efforts impose enormous stress on healthcare systems^6–8^.

Importantly, most HF cases are potentially preventable via aggressive control of risk factors^9^ which therefore represents a highly cost-effective approach^10^. As with any preventive measures, the early identification of individuals at risk is crucial as appreciated by recent HF guidelines, which emphasize the requirement for standardized screening strategies^11, 12^.

Current HF risk scores focus mainly on commonly acknowledged risk factors, e.g. including smoking history, systolic blood pressure and cholesterol levels. Several iterations of such clinical risk scores have been suggested in recent years, which often require the integration of data from heterogenous sources for acceptable performance^13^. Although most scores mainly comprise routinely available features, the combination of verbal interviews, physical measurements and clinical chemistry is time-consuming, expensive, and difficult to standardize.

Metabolites represent the end products of cellular processes and reflect the dynamic interactions between genes, environment and lifestyle factors. In recent years, proton nuclear magnetic resonance (^1^H-NMR) spectroscopy metabolomic profiling of serum samples has emerged as an effective assay to predict lifestyle^14–16^, disease onset and severity^17–23^, multimorbidity^24^, and all-cause mortality^25, 26^. Several single metabolites have been associated with incident HF^27, 28^. However, a systematic assessment and adequate benchmarking of the predictive value of serum metabolomics for incident HF has not been undertaken.

Here, we exploit the UK Biobank (UKB) resource to assess the suitability of serum metabolomics for HF risk stratification. Within UKB, large-scale metabolomic characterization (168 individual metabolites) via ^1^H-NMR spectroscopy has been conducted on ≈ 120,000 baseline serum samples collected between 2006 and 2010^19, 29^. Participants underwent extensive phenotypic characterization at enrolment and then health records follow-up to date. We first tested per-metabolite incident HF associations and describe a range of associated metabolites. Subsequently, elastic net (EN)-regularized COX proportional hazards (COX-PH) models were trained on 80 % of the dataset to generate risk scores relying on different combinations of age and sex, a state-of-the-art clinical HF risk prediction model (Pooled Cohort Equations to Prevent HF, PCP-HF), and the serum metabolome. We extensively assessed model performance and clinical utility on the remaining 20 % partition and find that metabolomics can enhance PCP-HF performance. Models solely relying on age, sex and metabolomics performed almost as well as PCP-HF, thus potentially displaying a promising alternative to more complex clinical scores. Our study applies machine learning to allow for minimising the number of measured features whilst preserving predictive performance.

## Methods

### Study design and population

The study was conducted within UKB, a large-scale prospective cohort representing the general UK population. 500,000 individuals were voluntarily enrolled between 2006-2010 (eligibility criteria: (I) aged 40-69 years at recruitment, (II) capacity to consent, (III) living within 20-25 miles of one of the assessment centres) via 22 assessment centres^30^ and participants received extensive baseline characterization via touchscreen questionnaires, verbal interviews, physical measurements, and the collection of relevant biological specimens. UKB regularly adds to their extensive resource via continued imaging programs, analysis of previously collected materials and ongoing clinical follow-up via electronic health records (Hospital Episode Statistics in England, Patient Episode Database for Wales, and Scottish Morbidity Record) and death register (NHS England and NHS Central Register) linkage.

Recently, UKB has incorporated serum ^1^H-NMR metabolomics on ≈ 120,000 non-fasting, venous serum samples of randomly selected study participants. This analysis was conducted in cooperation with Nightingale Health Plc, who provide a well-established ^1^H-NMR platform with broad regulatory approval^31^. Measured metabolites span routinely measured lipids as well as detailed lipoprotein subclass profiling, several amino acids, ketone bodies, glycolytic intermediates and markers of fluid balance and inflammation. We included all 168 original metabolite measurements.

This study included all UKB participants with non-missing values for the complete panel of 168 original ^1^H-NMR serum metabolite measurements in their initial assessment centre visit blood draw (55,253 female and 49,047 male participants, *n* = 104,300). We further excluded all participants with incomplete PCP-HF parameters (*n* = 17,654), those who already had or were considered at risk for HF (previous HF diagnosis, previous coronary artery disease (CAD) diagnosis or lipid-modifying pharmacotherapy; *n* = 15,842) or whose metabolomic measurements included data over 5 standard deviations (SD) from the mean (*n* = 2,493). In a final cohort of 68,311 patients, we investigated the association of the serum metabolome with incident HF. Subsequently, the cohort was split into derivation (80 %) and validation (20%) partitions stratified by HF. We trained EN models to predict incident HF risk on the derivation split, and subsequently assessed their predictive capacities in the validation partition (Figure 1).

**Figure 1:**
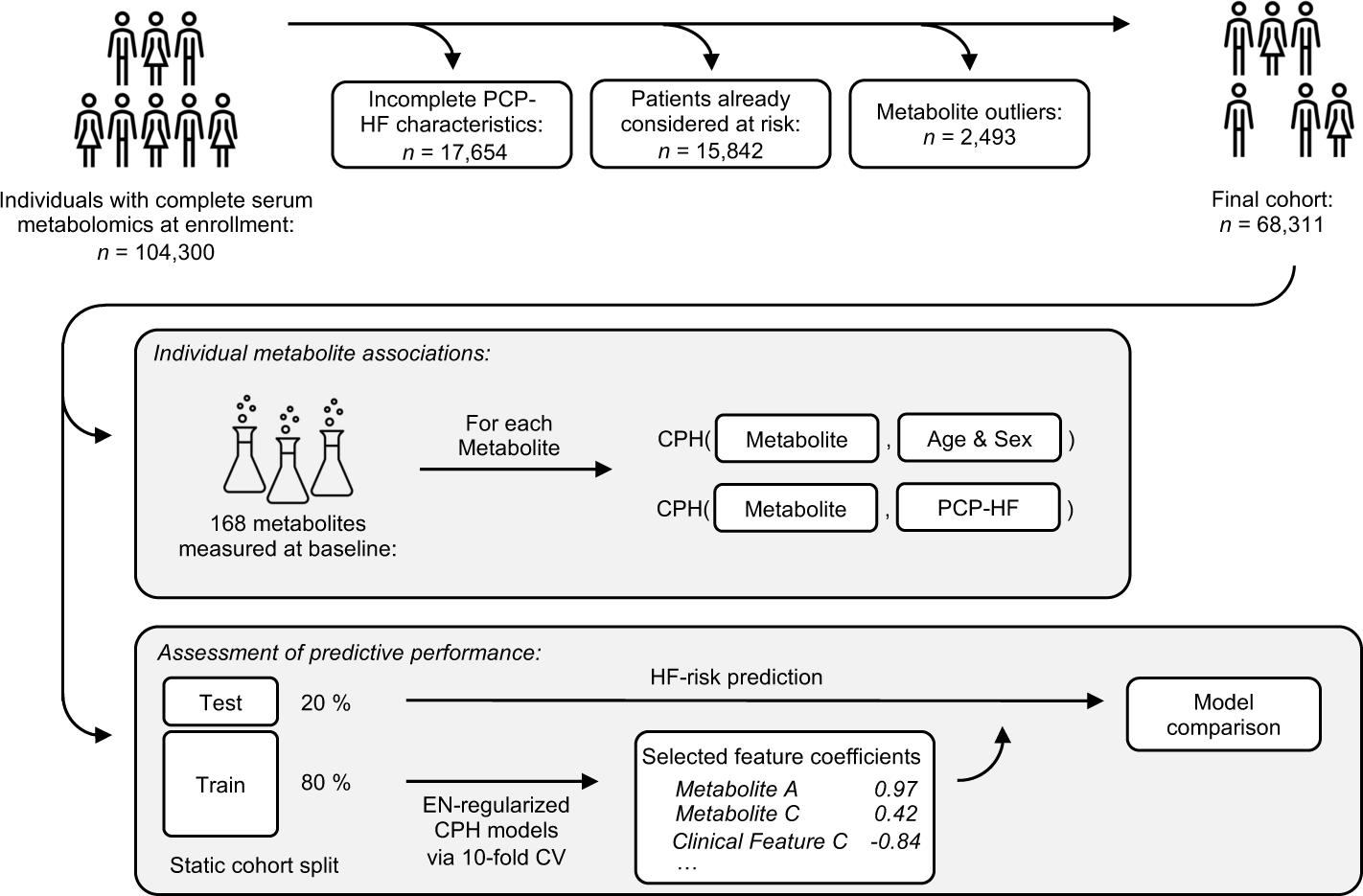
Study overview. After exclusion of individuals at risk, per-metabolite COX-PH models were fitted on the full cohort. Static cohort partition (80 % derivation, 20 % validation) allowed training and subsequent internal validation of risk prediction models. PCP-HF: Pooled Cohort Equations to Prevent HF, CPH: COX proportional hazards model, EN: elastic net, CV: cross-validation.

### Endpoint definition

Endpoint definitions were adapted as defined elsewhere^32, 33^. Onset of HF was defined as the earliest occurrence of (I) clinical diagnosis of HF (electronic health records), or (II) death by HF (death registers). Patients who self-reported HF in the baseline verbal interview were excluded from the study as having HF. We did not consider individuals with hypertrophic cardiomyopathy as having HF. Detailed definitions of all endpoints, including relevant UKB data fields and disease codes, can be found in Supplementary Table 1.

### Benchmark HF risk model and predictor extraction

PCP-HF is a well-validated model for 10-year HF risk, which has been developed on 7 smaller, community-based U.S. cohorts^1^. PCP-HF includes basic sociodemographic parameters (age, race, sex), patient history (current smoking, hypertension treatment, diabetes treatment), physical measurements (systolic blood pressure, body mass index, QRS duration), and clinical chemistry (total cholesterol, high-density lipoprotein cholesterol (HDL-C)). All PCP-HF components except QRS duration (due to a lack of individuals that underwent ECG within UKB) were extracted from the UKB dataset. PCP-HF weighs QRS duration relatively weakly (original average risk equation coefficients: QRS duration: 0.92, age: 29.28, BMI: 15.23) and has been validated externally by the authors without the inclusion of QRS duration^34^. Clinical chemistry parameters were derived from non-NMR measurements that had been conducted at a central UKB laboratory between 2014 and 2017^35^. Systolic blood pressure was routinely measured twice a few moments apart; we selected the lower reading. Where automated blood pressure readings were faulty manual measurements were conducted, which have been used by us in such cases. All relevant data fields and corresponding disease/medication codes can be found in Supplementary Table 2.

### Survival analysis for individual metabolite associations

All metabolites were log-normalized and standardized to a mean of 0 and a standard deviation of 1. We assessed incident HF via fitting of COX-PH models for each individual metabolite (conducted using the *survival* package in R). Models were adjusted for age and sex or the complete panel of PCP-HF characteristics (also including age and sex; Supplementary Tables 3 and 4). P-values were adjusted for multiple testing using the Benjamini-Hochberg method.

### Elastic net model generation and evaluation

To select metabolites with the highest predictive capacities, we trained EN-regularized COX-PH models on the derivation cohort (baseline characteristics in Supplementary Table 5). Models were trained using an internal to the derivation cohort 10-fold cross-validation approach and model hyperparameters were optimized for discriminative performance (Harrel’s C; most regularized model within one standard error of maximum performance; conducted using the *hdnom* package). All model coefficients and hyperparameters can be found in Supplementary Tables 6 and 7. Using the fitted model coefficients, we calculated individual HF risk for the validation cohort. We assessed discriminative capacities via calculation of Harrel’s C, as well as sensitivity and specificity at the Youden point. We implemented comparative metrics and calculated case, non-case and overall continuous net reclassification improvement (NRI) as well as absolute and relative integrated discrimination improvement (IDI) (performance within the derivation cohort is reported in Supplementary Tables 8 and 9). Receiver operator characteristic (ROC) curves were generated using the *pROC* package, decision curve analysis (DCA) plots using the *dca* package on the calibration cohort (see below). Individuals were grouped into risk quintiles according to predicted HF risk. Both incidence and survival were plotted according to risk quintile. Calibration plots of absolute predicted vs. observed 10-year HF risk were generated on all individuals that had not been censored prior to 10 years of follow-up for reasons other than the development of HF. Model feature Spearman correlations were calculated and plotted using the *corrplot* package. Network visualization followed a standard weighted gene co-expression network analysis workflow^36^. Metabolite correlations were calculated and transformed into an adjacency matrix using a soft thresholding power of β = 30 (chosen via visual inspection of scale-free topology fit index R^2^ and mean connectivity). Network edges were dichotomized via hard thresholding (threshold = 0.2) of the subsequently generated topological overlap matrix to generate an unweighted network view. Metabolite nodes were coloured in saturated colours if they were represented in the final PCP-HF + Metabolomics model and shrunken according to the degree of connectivity (higher connectivity – smaller size). Network generation and visualization were conducted using the *WGCNA*^37^ and the *igraph* packages.

### Significance, software, and code availability

All analyses were conducted in R (v.4.2.3). Statistical significance was false discovery rate (FDR)-controlled using the Benjamini-Hochberg method as appropriate, significance was defined as adjusted P < 0.01. The full code will be available upon publication.

### Data availability

UKB’s data is publicly available to approved researchers at https://www.ukbiobank.ac.uk/enable-your-research. UKB data was accessed under application ID 98729. Detailed endpoint and predictor definitions can be found in Supplementary Tables 1 and 2.

## Results

### Study cohort

Amongst the eligible study population, the median age at recruitment was 57 years (IQR 49 - 62) and 55.84 % were female. Individuals who would eventually develop HF (n = 1,460 (2.14 %)) were more likely to be older, male, and exhibited a range of different baseline characteristics and incident endpoint frequencies (Table 1). This included highly significant differences for several sociodemographic and lifestyle factors (age, sex, education, smoking status and alcohol consumption), clinical chemistry measurements (HDL cholesterol, triglycerides, glucose, glycated haemoglobin, creatinine and C-reactive protein), physical measurements (systolic blood pressure, waist-to-hip ratio and BMI) and the incidence of hypertension and CAD. All significant changes followed the expected direction.

**Table 1:**
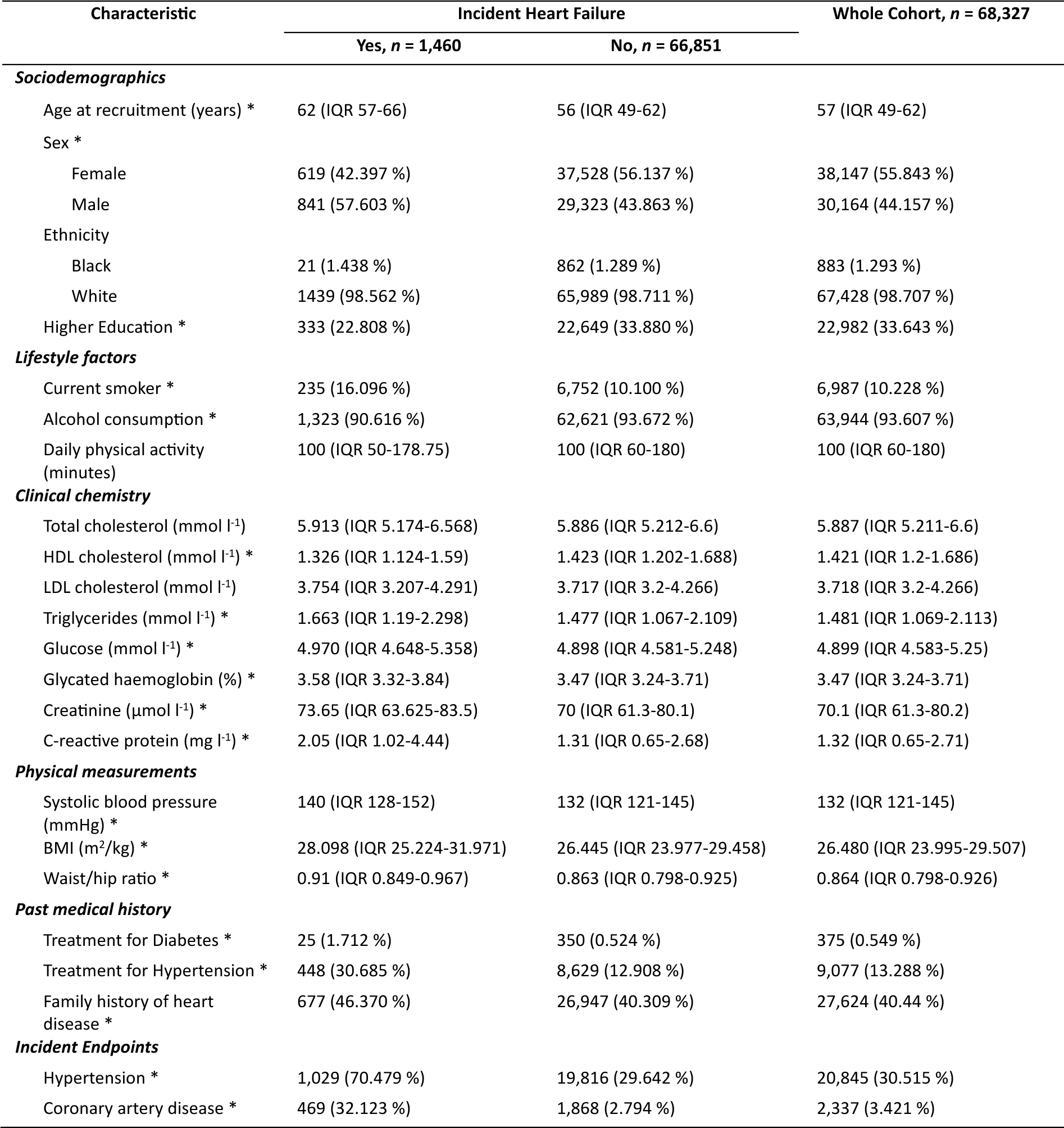
Population characteristics at baseline and incident endpoints. Listed clinical chemistry measurements were not ^1^H-NMR-derived. Median (IQR) for continuous, *n* (%) for categorical variables. Mann-Whitney-U for continuous, Chi-Square for categorical variables; BH-correction for multiple testing. * P < 0.01.

| Characteristic | Incident Heart Failure |  | Whole Cohort, *n* = 68,327 |
| --- | --- | --- | --- |
|  | Yes, *n* = 1,460 | No, *n* = 66,851 |  |
| ***Sociodemographics*** |  |  |  |
| Age at recruitment (years) * | 62 (IQR 57-66) | 56 (IQR 49-62) | 57 (IQR 49-62) |
| Sex * |  |  |  |
| Female | 619 (42.397 %) | 37,528 (56.137 %) | 38,147 (55.843 %) |
| Male | 841 (57.603 %) | 29,323 (43.863 %) | 30,164 (44.157 %) |
| Ethnicity |  |  |  |
| Black | 21 (1.438 %) | 862 (1.289 %) | 883 (1.293 %) |
| White | 1439 (98.562 %) | 65,989 (98.711 %) | 67,428 (98.707 %) |
| Higher Education * | 333 (22.808 %) | 22,649 (33.880 %) | 22,982 (33.643 %) |
| ***Lifestyle factors*** |  |  |  |
| Current smoker * | 235 (16.096 %) | 6,752 (10.100 %) | 6,987 (10.228 %) |
| Alcohol consumption * | 1,323 (90.616 %) | 62,621 (93.672 %) | 63,944 (93.607 %) |
| Daily physical activity (minutes) | 100 (IQR 50-178.75) | 100 (IQR 60-180) | 100 (IQR 60-180) |
| ***Clinical chemistry*** |  |  |  |
| Total cholesterol (mmol l<sup>-1</sup>) | 5.913 (IQR 5.174-6.568) | 5.886 (IQR 5.212-6.6) | 5.887 (IQR 5.211-6.6) |
| HDL cholesterol (mmol l<sup>-1</sup>) * | 1.326 (IQR 1.124-1.59) | 1.423 (IQR 1.202-1.688) | 1.421 (IQR 1.2-1.686) |
| LDL cholesterol (mmol l<sup>-1</sup>) | 3.754 (IQR 3.207-4.291) | 3.717 (IQR 3.2-4.266) | 3.718 (IQR 3.2-4.266) |
| Triglycerides (mmol l<sup>-1</sup>) * | 1.663 (IQR 1.19-2.298) | 1.477 (IQR 1.067-2.109) | 1.481 (IQR 1.069-2.113) |
| Glucose (mmol l<sup>-1</sup>) * | 4.970 (IQR 4.648-5.358) | 4.898 (IQR 4.581-5.248) | 4.899 (IQR 4.583-5.25) |
| Glycated haemoglobin (%) * | 3.58 (IQR 3.32-3.84) | 3.47 (IQR 3.24-3.71) | 3.47 (IQR 3.24-3.71) |
| Creatinine (μmol l<sup>-1</sup>) * | 73.65 (IQR 63.625-83.5) | 70 (IQR 61.3-80.1) | 70.1 (IQR 61.3-80.2) |
| C-reactive protein (mg l<sup>-1</sup>) * | 2.05 (IQR 1.02-4.44) | 1.31 (IQR 0.65-2.68) | 1.32 (IQR 0.65-2.71) |
| ***Physical measurements*** |  |  |  |
| Systolic blood pressure (mmHg) * | 140 (IQR 128-152) | 132 (IQR 121-145) | 132 (IQR 121-145) |
| BMI (m<sup>2</sup>/kg) * | 28.098 (IQR 25.224-31.971) | 26.445 (IQR 23.977-29.458) | 26.480 (IQR 23.995-29.507) |
| Waist/hip ratio * | 0.91 (IQR 0.849-0.967) | 0.863 (IQR 0.798-0.925) | 0.864 (IQR 0.798-0.926) |
| ***Past medical history*** |  |  |  |
| Treatment for Diabetes * | 25 (1.712 %) | 350 (0.524 %) | 375 (0.549 %) |
| Treatment for Hypertension * | 448 (30.685 %) | 8,629 (12.908 %) | 9,077 (13.288 %) |
| Family history of heart disease * | 677 (46.370 %) | 26,947 (40.309 %) | 27,624 (40.44 %) |
| ***Incident Endpoints*** |  |  |  |
| Hypertension * | 1,029 (70.479 %) | 19,816 (29.642 %) | 20,845 (30.515 %) |
| Coronary artery disease * | 469 (32.123 %) | 1,868 (2.794 %) | 2,337 (3.421 %) |

### Several individual metabolites are associated with incident HF

Individual metabolite associations with incident HF were assessed via per-metabolite fitting of COX-PH models. Models adjusted for age and sex revealed 90/168 (54 %)) significant metabolite associations. Fewer significant metabolite associations (48/168 (29 %)) were observed in models more extensively adjusted for all PCP-HF features. The overlap of significant metabolites was 25/168 (15 %) (detailed statistics for all metabolites in Supplementary Table 4).

Within age- and sex-adjusted models (Figures 2A and B), we observed consistent association patterns for several lipoprotein subspecies. The concentration of very low-density lipoprotein (VLDL) particles, as well as their (free/esterified) cholesterol, phospholipid, triglyceride and total lipid contents, were positively associated with incident HF. Inverse associations were observed for HDL and, to a lesser extent, low-density lipoprotein (LDL) subspecies. Numerous additional metabolites were significantly associated with incident HF, including predominantly positive correlations of different ketone bodies, glycolytic metabolites, and glycoprotein acetyls. Within the measured amino acids, higher levels of alanine, glutamine, glycine and histidine were negatively associated with incident HF, whilst phenylalanine and tyrosine showed a positive association. Levels of apolipoproteins, lipoprotein sizes, total cholines and phosphatidylcholines, phosphoglycerides and sphingomyelins showed significant negative associations. The degree of fatty acid (FA) saturation influenced incident HF association; saturated and monounsaturated FAs were found to be positively associated, whilst levels of all other FAs including the degree of FA unsaturation were negatively associated with incident HF. Significant inverse associations with incident HF were found for albumin.

**Figure 2:**
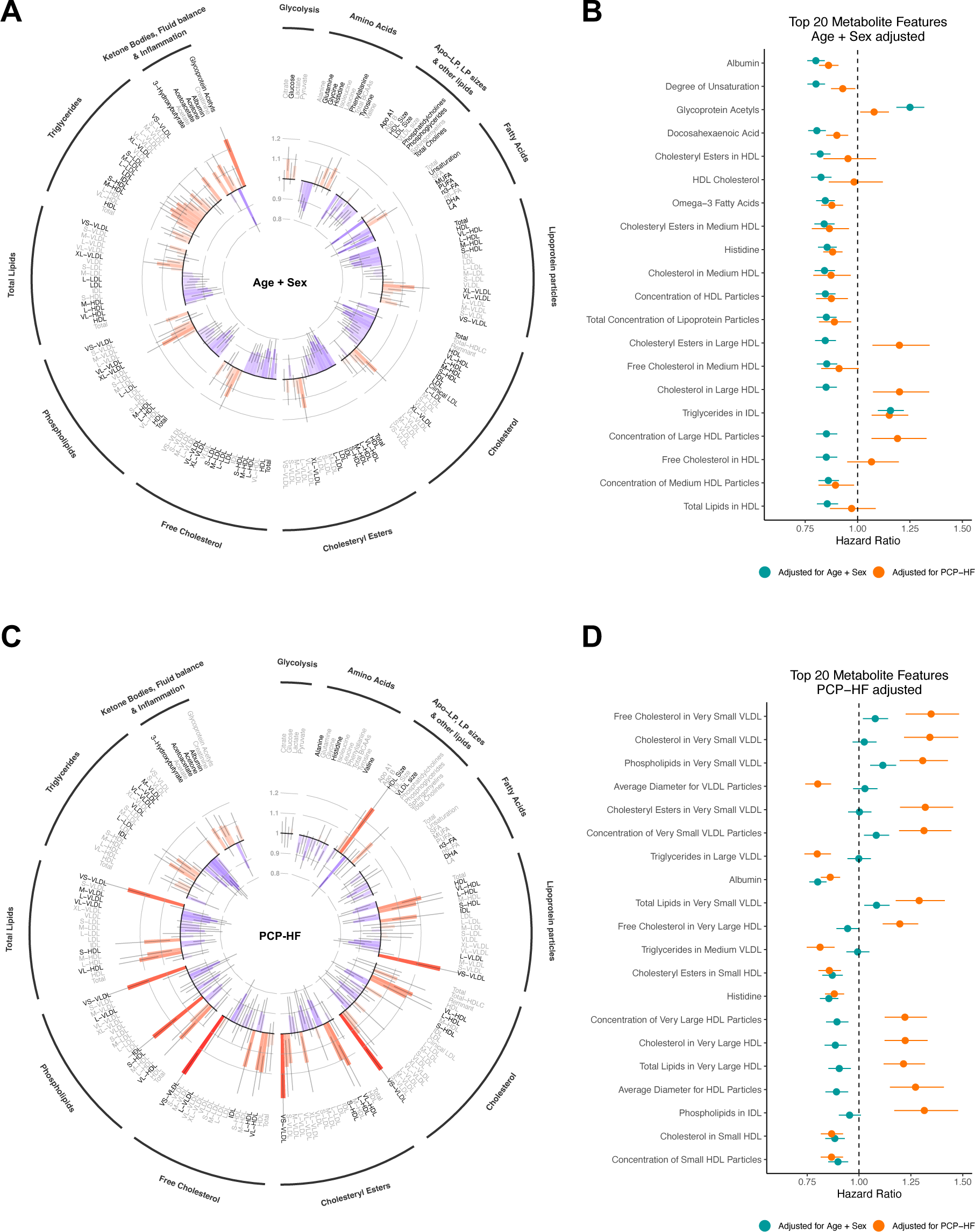
Individual metabolite associations. Per-metabolite COX-PH models adjusted for age and sex (A), or PCP-HF features (C) reveal several metabolites significantly associated (FDR-controlled P < 0.01, dark grey labels) with incident HF. Bar colours illustrate the directionality of associations: positive (red), negative (blue). Forrest plots (B and D) illustrate hazard ratios of the 20 most significant features (sorted by P, in descending order; green: adjusted for age + sex, orange: adjusted for PCP-HF). PCP-HF: Pooled Cohort Equations to Prevent HF.

Following adjustment for PCP-HF (Figures 2C and D), we observed more heterogenous metabolite association patterns within metabolite groups. Amongst different lipoprotein subclasses, VLDL and HDL particles showed the most significant associations with HF. We observed that larger HDL particles were positively associated with HF, whilst smaller VLDL subspecies showed inverse associations. These effects were supported by the associations of lipoprotein particle sizes (significantly positive for HDL and negative for VLDL). Similar to age- and sex-adjusted models, (free/esterified) cholesterol, phospholipid, triglyceride and total lipid content of the different lipoprotein particles correlated well with the associations of their respective concentrations. Ketone bodies and glycoprotein acetyls were positively associated with incident HF. Albumin, as well as all significantly associated amino acids, were inversely associated with HF. The concentrations of omega-3 FAs and docosahexaenoic acid were found to be negatively associated with incident HF. No glycolytic metabolite reached the significance threshold.

Concentration of albumin (HR = 0.800 (0.760 - 0.843), P = 8.5 × 10^−15^), the degree of unsaturation of FAs (HR = 0.801 (0.761 - 0.844), P = 8.5 × 10^−15^) and glycoprotein acetyls (HR = 1.250 (1.186 – 1.318), P = 8.7 × 10^−15^) were most significantly associated with incident HF in age and sex adjusted models. Following adjustment for PCP-HF characteristics, the most significant associations were observed for free cholesterol (HR = 1.348 (1.225 – 1.482), P = 1.349 × 10^−7^), cholesterol (HR = 1.342 (1.217 – 1.479), P = 1.934 × 10^−7^), and phospholipid (HR = 1.307 (1.196 – 1.429), P = 1.934 × 10^−7^) content of VLDLs.

### Utilization of serum metabolomics improves incident HF risk discrimination

Our dataset was split in a derivation (80%) and validation (20%) cohort stratified by HF prevalence. Patient baseline characteristics and incident endpoints were well-balanced between the two partitions (Supplementary Table 5). We fitted EN-regularized COX-PH models for either age and sex or the complete PCP-HF set on the derivation cohort, both with and without the addition of serum metabolomics. Final models retained age and sex, 9/10 (PCP-HF model) and 7/10 (PCP-HF + Metabolomics model) additional clinical features as well as 21/168 (Age & Sex + Metabolomics model) and 16/168 (PCP-HF + Metabolomics model) metabolites (Supplementary Tables 6 and 7, including individual feature coefficients and optimized model hyperparameters).

Model performance was evaluated on the validation cohort (*n* = 13,662; ≈ 170,000 person-years of follow-up). At various risk thresholds, ROC and DCA curves of models that integrated metabolomics showed superior performance over their respective non-metabolomic benchmarks (Figure 3). Of note, the combination of age, sex and metabolomics and PCP-HF exhibited undulating curves, with both respective models performing better within certain threshold ranges. To assess model performance, we calculated Harrel’s C, sensitivity, and specificity at Youden. We compared models via calculation of continuous NRI and IDI. Generally, we found that models trained under inclusion of metabolomic profiles possessed improved discriminatory capacities (Age & Sex vs. Age & Sex + Metabolomics: ΔC = 0.044 (P = 1.741 × 10^−17^ amongst derivation cross-validation splits), NRI = 0.667; PCP-HF vs. PCP-HF + Metabolomics: ΔC = 0.013 (P = 3.664 × 10^−8^ amongst derivation cross-validation splits), NRI = 0.287; Tables 2 and 3). In line with these findings, we consistently observed positive IDIs when adding metabolomic features (Table 3). The comprehensive PCP-HF model showed only modest reclassification improvements over the model including age, sex, and metabolomics (ΔC = 0.010 (P = 2.451 × 10^−8^ amongst derivation cross-validation splits), NRI = 0.097, relative IDI = 13.445 %).

**Figure 3:**
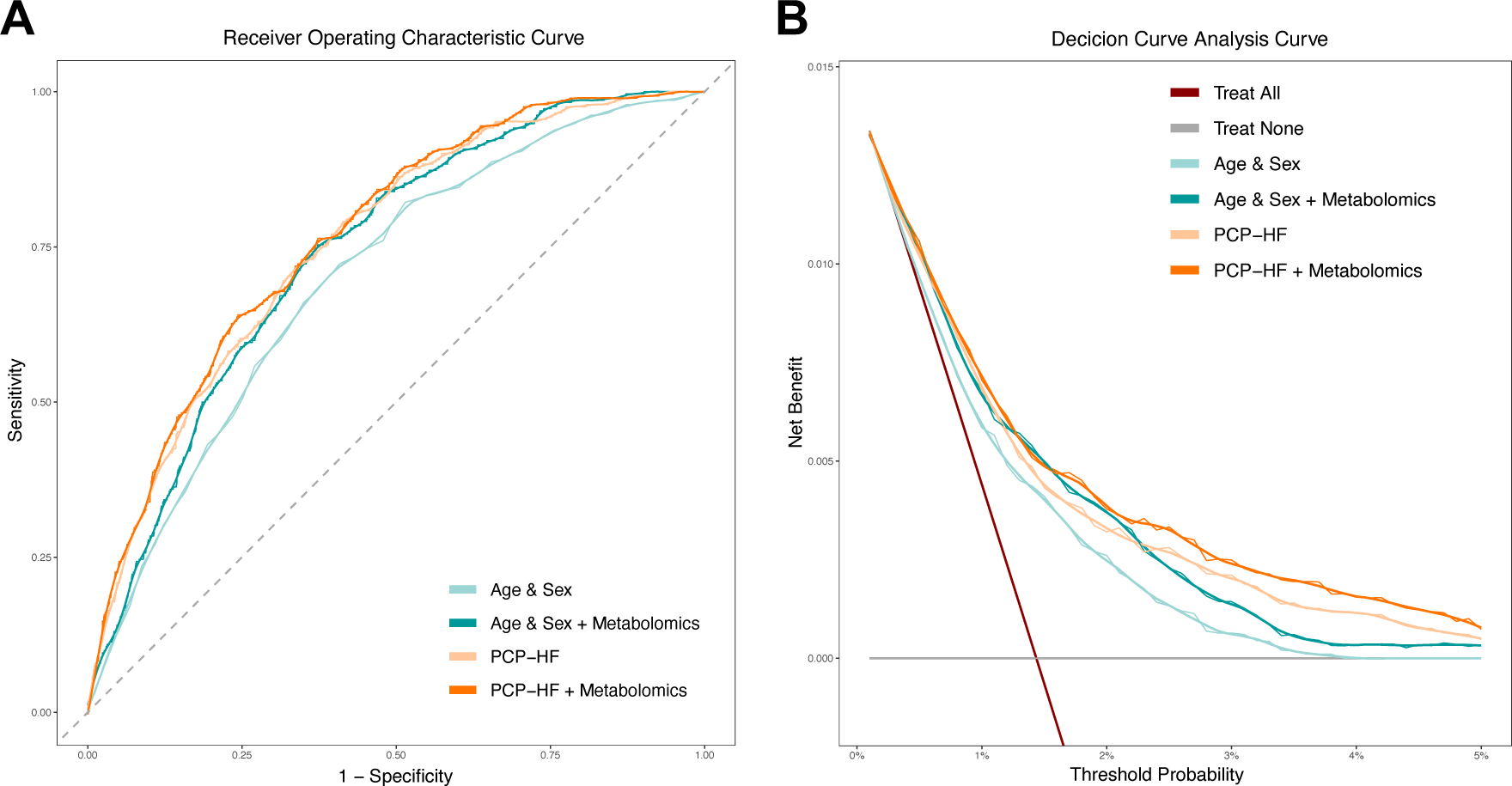
Internal model validation. Receiver operator characteristic (ROC; A) and decision curve analysis (DCA; B) curves illustrate discriminative performance of the different models. Smoothened curves (bold) are superimposed on individual data points (fine). PCP-HF: Pooled Cohort Equations to Prevent HF.

**Table 2:** Internal model validation: absolute discriminative capacities. Model performance is assessed in the validation partition. Sensitivity and Specificity are calculated at the Youden point. PCP-HF: Pooled Cohort Equations to Prevent HF.

|  | Age & Sex | Age & Sex<br>+ Metabolomics | PCP-HF | PCP-HF<br>+ Metabolomics |
| --- | --- | --- | --- | --- |
| Harrell's C | 0.701 | 0.744 | 0.755 | 0.768 |
| Sensitivity | 0.723 | 0.753 | 0.719 | 0.640 |
| Specificity | 0.592 | 0.627 | 0.664 | 0.757 |

**Table 3:**
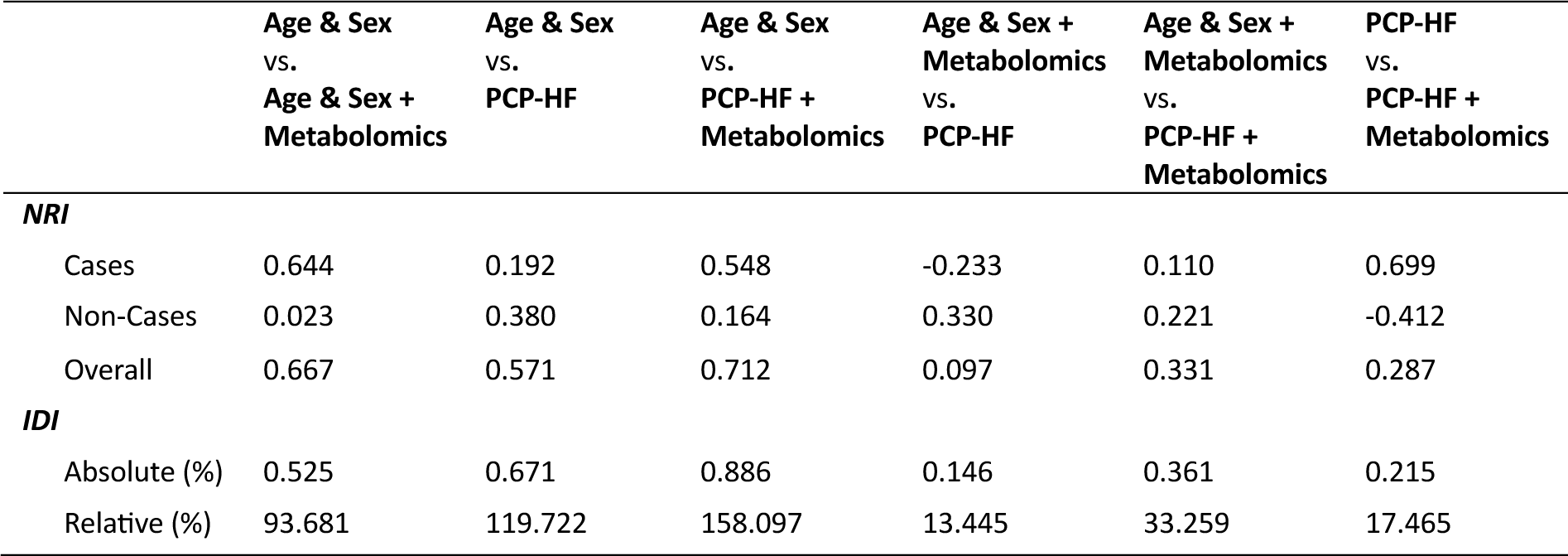
Internal model validation: metrics of relative performance. Model performance is assessed on the validation partition. NRI: Net reclassification improvement, IDI: Integrated discrimination improvement. PCP-HF: Pooled Cohort Equations to Prevent HF.

|  | Age & Sex<br>vs.<br>Age & Sex +<br>Metabolomics | Age & Sex<br>vs.<br>PCP-HF | Age & Sex<br>vs.<br>PCP-HF +<br>Metabolomics | Age & Sex +<br>Metabolomics<br>vs.<br>PCP-HF | Age & Sex +<br>Metabolomics<br>vs.<br>PCP-HF +<br>Metabolomics | PCP-HF<br>vs.<br>PCP-HF +<br>Metabolomics |
| --- | --- | --- | --- | --- | --- | --- |
| **NRI** |  |  |  |  |  |  |
| Cases | 0.644 | 0.192 | 0.548 | -0.233 | 0.110 | 0.699 |
| Non-Cases | 0.023 | 0.380 | 0.164 | 0.330 | 0.221 | -0.412 |
| Overall | 0.667 | 0.571 | 0.712 | 0.097 | 0.331 | 0.287 |
| **IDI** |  |  |  |  |  |  |
| Absolute (%) | 0.525 | 0.671 | 0.886 | 0.146 | 0.361 | 0.215 |
| Relative (%) | 93.681 | 119.722 | 158.097 | 13.445 | 33.259 | 17.465 |

### Metabolomics facilitate accurate HF risk prediction

To further explore the translational potential of our risk models we assessed cumulative HF incidence, model calibration and HF-free survival according to predicted risk. Stratification of HF incidence amongst risk quintiles was improved following the addition of serum metabolomics (Incidence_HF, top risk quintile_: Age & Sex = 4.61 %, Age & Sex + Metabolomics = 5.38 %, PCP-HF = 5.56 %, PCP-HF + Metabolomics = 5.78 %; Incidence_HF, bottom risk quintile_: Age & Sex = 0.55 %, Age & Sex + Metabolomics = 0.15 %, PCP-HF = 0.26 %, PCP-HF + Metabolomics = 0.11 %; Figure 4A). The largest differences in HF incidence amongst risk quintiles were observed using the combination of clinical characteristics and metabolomics (ΔIncidence_HF, top vs. bottom risk quintile_: 5.67 %; Figure 4A). Model calibration was assessed on all individuals who had completed 10 years of follow-up or developed HF beforehand. We observed fair, highly similar calibration in all models except the more inaccurate model based solely on age and sex (Figure 4B). Finally, Kaplan-Meier plots highlight survival stratification for the complete follow-up period, which appears superior in more extensive models including both clinical PCP-HF characteristics and metabolite features (Figure 4C). In summary, we observed marked improvements in all models over the very basic age and sex model. Notably, the simple addition of metabolomics to such models closely resembles the more extensive PCP-HF model.

**Figure 4:**
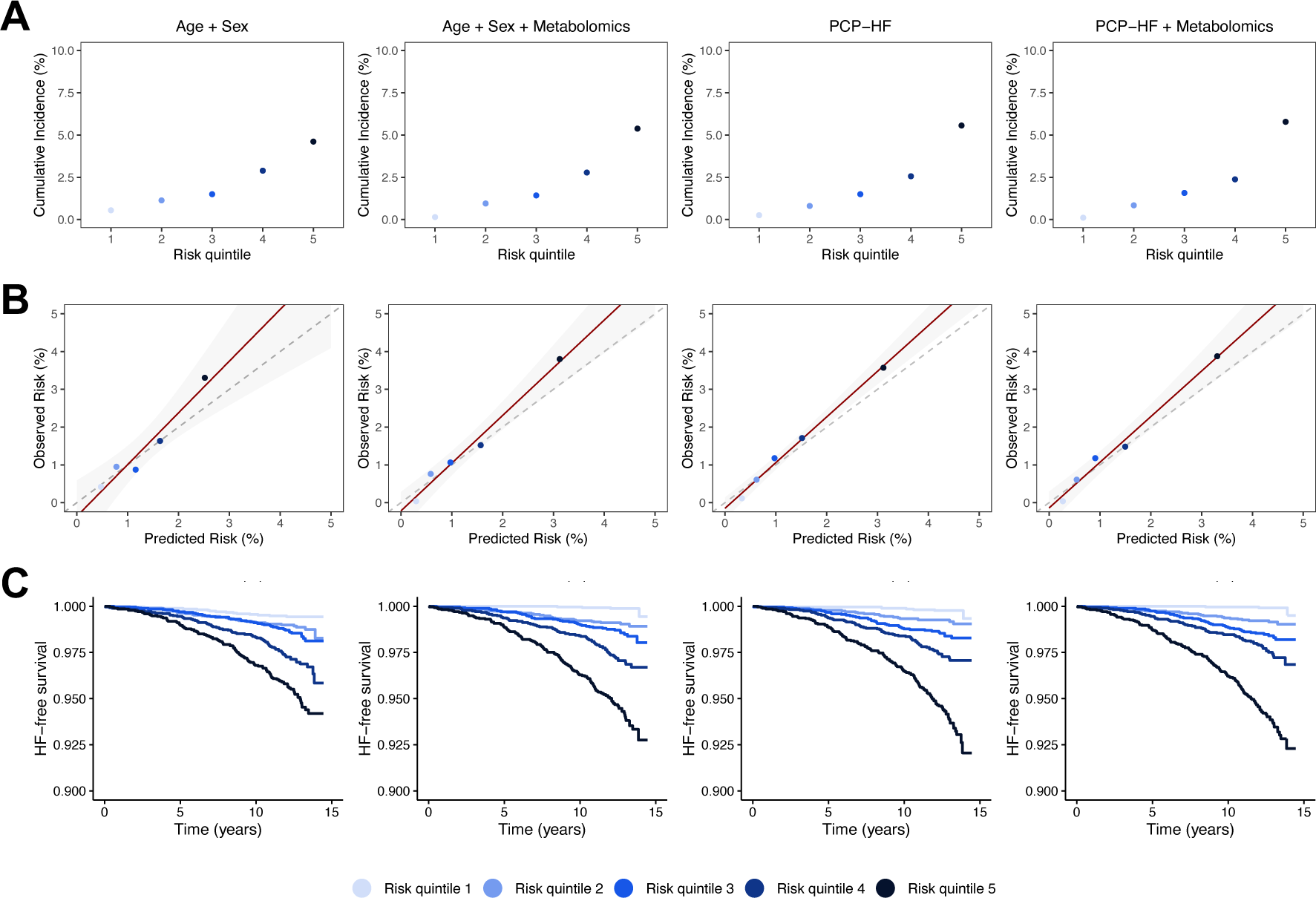
Metabolomics improve risk and survival stratification. Cumulative HF incidence (A), model calibration (observed vs. predicted incident HF risk; B) and HF-free survival (C) according to risk quintile (blue shades). Calibration was assessed on a follow-up timeframe of 10 years; all individuals who were censored prior to 10 years for reasons other than the development of HF were excluded. PCP-HF: Pooled Cohort Equations to Prevent HF.

### Effective feature reduction using elastic net models

Reducing the number of measured features is essential to provide preventive screening in a cost-efficient manner. In our work, the selection of features containing most predictive information is achieved via EN-regularized COX regression models, which were trained in a 10-fold cross-validation approach.

Figure 5A demonstrates the poor correlation of retained features. Network visualization of the measured metabolites highlights the effective representation of highly connected clusters by single metabolites whilst several, uncorrelated metabolite features were included in the final model (Figure 5B). The retained metabolites included five amino acids, four lipoprotein subspecies, three ketones, glucose, omega-3 FAs, glycoprotein acetyls and albumin (model coefficients of all clinical and metabolite features in Supplementary Table 6).

**Figure 5:**
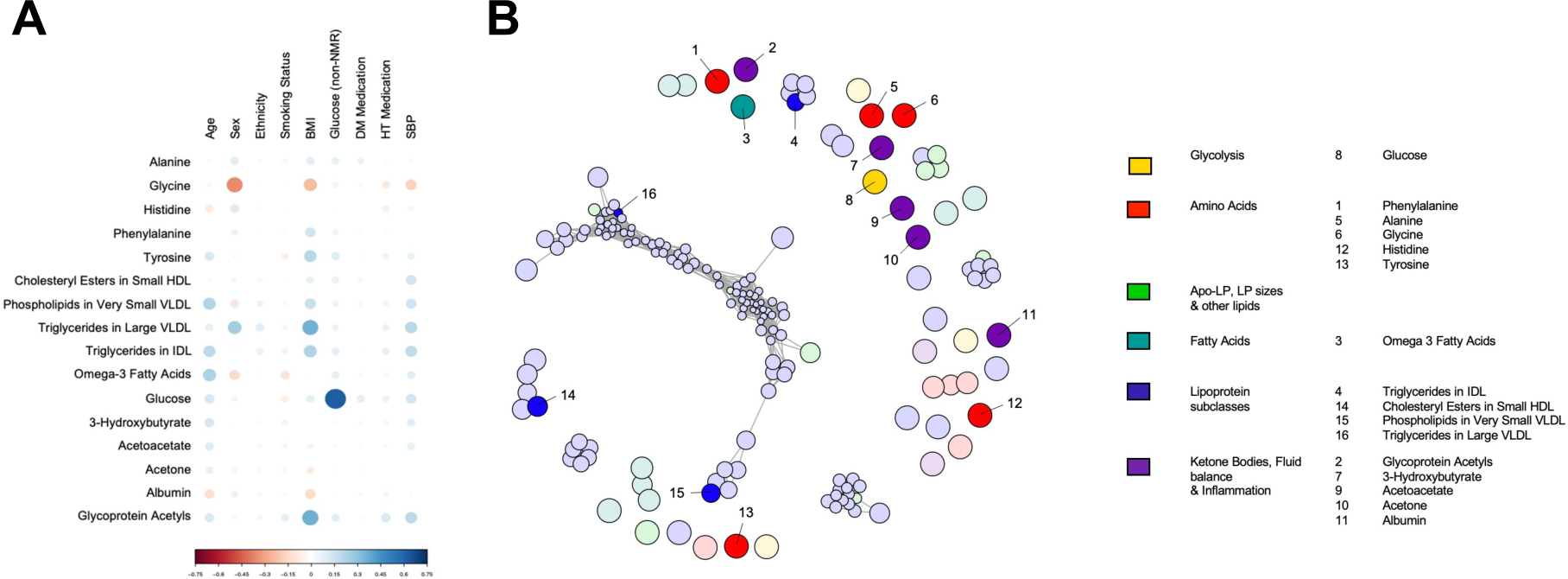
Metabolic risk is effectively represented by key features. Correlation plot of retained clinical and metabolite features of the most comprehensive PCP-HF + Metabolomics model (A) and network visualization of the measured metabolome (B). Network edges represent adjacency > 0.2. Nodes are highlighted in saturated colours if they were used by the final model. Node size is shrunk according to node connectivity.

## Discussion

The current lifetime prevalence of HF is approximately 20%^8^. A large proportion of these cases could be prevented^9^. Improved risk stratification is vital for the effective implementation of preventive strategies. Several incident HF risk prediction models have been suggested in recent years but none has yet been clinically adopted. Although many approaches achieve good discriminative performance, available models come with limitations such as derivation from inadequately sized, nonrepresentative cohorts or the integration of heterogenous and poorly standardizable data sources^13^.

HF is well-described to have a strong metabolic basis. This applies both locally, where cardiac metabolism plays a central role in cardiac remodelling processes^38^, and systemically, where hemodynamic changes potently induce metabolic alterations in several organs^39^. Moreover, several HF risk factors are associated with broad metabolic perturbations^40^. Measuring the blood metabolome thus has rich potential to allow versatile inferences on cardiac pathophysiology and risk profile via a single blood draw.

Within this study comprising 68,311 individuals, > 0.8 million person-years of follow up and 1,460 HF cases, we highlight the value of serum metabolomics for the prediction of incident HF. Our results reveal a range of incident HF - associated metabolites, even after adjustment for clinical HF risk. Moreover, we show that the serum metabolome can be leveraged to improve state-of-the-art clinical risk prediction models. Narrow models relying only on a combination of age, sex and metabolomics perform as well as more complex models that incorporate multiple clinical parameters.

Our finding of association of incident HF with individual metabolites includes overlap with previously reported associations and accepted pathomechanisms. For example, high levels of atherogenic VLDL (subspecies) increased the risk of incident HF. The concentrations of HDL and its subspecies, possessing anti-atherogenic properties, were found to be inversely associated with incident HF. The positive association of ketone bodies (which are commonly elevated in the serum of HF patients and used as a cardiac fuel source in such settings^41^), glycolytic metabolites (also increasingly utilized by the failing heart^42^ and closely associated with metabolic risk factors^43^), and glycoprotein acetyls (reflective of detrimental systemic inflammation^44, 45^) were also not unexpected. Individuals with higher levels of amino acids and albumin were less likely to develop HF, consistent with the common observation of decreased levels of those metabolites in settings of increased cardiac workload^46, 47^. In line with previously published results from PROSPER and FINRISK^27^, tyrosine and its precursor phenylalanine were positively associated with incident HF, in contrast to all other amino acids. This phenomenon has been repeatedly observed and extensively discussed in the context of altered protein uptake and turnover, impaired renal clearance or the utilisation of those amino acids for the synthesis of catecholamines^27, 48, 49^. The degree of FA unsaturation was inversely associated with HF, thus likely reflecting underlying HF risk factors that arise from the participant’s diet and lifestyle^14^. In alignment with previous studies, we demonstrate that ^1^H-NMR serum metabolomics are well-suited to consistently capture common metabolic signatures of cardiometabolic risk factors, cardiac remodelling and HF.

Importantly, several metabolite associations were retained even after adjustment for clinical HF risk, thus suggesting that the serum metabolome contains information on HF risk that is not effectively captured by clinical risk scores. We therefore assessed whether we could improve on existing risk stratification tools via the integration of ^1^H-NMR metabolomics. Previous studies have convincingly shown that metabolomics provide additive predictive value over clinical risk stratification for a multi-disease spectrum^17, 18, 23^. Our results, benchmarking against a HF-specific risk score and specifically excluding high-risk patients, align with such results as they demonstrate improved discriminative performance upon consideration of serum metabolomics.

The performance of relatively simple models only including age, sex and the plasma metabolome was strikingly good, in some indices even outperforming PCP-HF (consistent amongst derivation and validation partitions). From our perspective, a single-domain assay such as ^1^H-NMR metabolomics offers several advantages over the limitations that come with currently available risk prediction models: (I) Clinical risk scores often strongly focus on commonly acknowledged high-risk factors (e. g. history of valvular heart disease, previous diagnosis of CAD). Such models might therefore exhibit excellent discriminatory capacities but preferentially select subpopulations who were already considered at risk or are even already receiving targeted preventive measures^50^.Individuals without obvious risk constellations might particularly benefit from targeted screening strategies. With the exception of hypertensive participants (hypertensive treatment is considered by PCP-HF), our models were trained and evaluated on such patients that might have otherwise been overlooked. Moreover, (II) assays for ^1^H-NMR serum metabolomics are highly reliable and inexpensive^31, 51^. The methodology enables capturing via a single blood draw (as already collected in many countries as part of routine health checks) of extensive systemic information that might otherwise require interviews, clinical chemistry, physical measurements and imaging. Such heterogenous data sources are difficult to standardize, expensive to collect, and less effectively scalable. In addition, (III) as shown in recent approaches^17, 18, 23^, serum metabolomics simultaneously inform on multi-disease risk for several entities other than HF. Whilst disease-specific benchmarking against the clinical state-of-the-art is warranted, simultaneous multi-disease prediction might further expand the utility of serum metabolomics.

The strengths of our study are its large sample size and the exclusion of patients already known to be at risk, thereby also avoiding treatment-associated biases that could arise from e. g. lipid-modifying therapy. We benchmarked for disease-specific, validated clinical risk scores and used a well-established metabolomic platform that has received regulatory approval^19^. However, several challenges remain before clinical implementation. The UKB population is known to be not fully representative, with generally older and healthier individuals compared to the sampling population (“healthy volunteer” bias^52^). Moreover, predictor variable collection was highly standardized within UKB, which might not be directly transferable to a healthcare system level. Although we confirmed internal validity, our results would still benefit from external validation in a different population. Plasma samples were taken in a non-fasting state, which might lead to some variability. Finally, the metabolomic assay that was employed provides measurements for a wide range of lipids and lipid subspecies but is relatively restricted for other low-molecular weight metabolites.

In conclusion, we demonstrate that ^1^H-NMR serum metabolomics could potentially be effectively implemented as a single-domain screening tool for incident HF risk. We demonstrate that machine learning can be effectively applied to reduce the number of features used for risk prediction (80-90 % feature reduction) whilst simultaneously preserving predictive performance. In the context of HF, several large metabolite clusters were effectively represented by single key metabolite features with high predictive value. This generates overt potential for a cost-effective implementation, thus potentially facilitating clinical adoption.

## Data Availability

UKB data is publicly available to approved researchers at https://www.ukbiobank.ac.uk/enable-your-research. UKB data was accessed under application ID 98729. Detailed endpoint and predictor definitions can be found in Supplementary Tables 1 and 2.

## Acknowledgements

RO is supported by a British Heart Foundation (BHF) PhD studentship (FS/19/58/34895). KT is supported by a BHF project grant (PG/20/10387). AMS is supported by the BHF (CH/1999001/11735, RG/20/3/34823, RE/18/2/34213).

