## Supplementary Tables for "Serum metabolomics improve risk stratification for incident heart failure"

| Field names | Data fields | Data coding | Meaning |
|---|---|---|---|
| ***Coronary artery disease*** | | | |
| Non-cancer Illness Codes | 20002 | 1075 | Heart attack/myocardial infarction |
| Operation code | 20004 | 1070, 1095 | Coronary angioplasty ± stent, Coronary artery bypass grafts |
| Underlying (primary) cause of death: ICD10 | 40001 | I21.X, I21.0, I21.1, I21.2, I21.3, I21.4, I21.9;<br>I22.X, I22.0, I22.1, I22.8, I22.9;<br>I23.X, I23.1, I23.2, I23.3, I23.6, I23.8;<br>I24.1;<br>I25.2 | Acute myocardial infarction; Subsequent myocardial infarction; Certain current complications following acute myocardial infarction; Dressler's syndrome; Old myocardial infarction |
| Contributory (secondary) causes of death: ICD10 | 40002 | I21.X, I21.0, I21.1, I21.2, I21.3, I21.4, I21.9;<br>I22.X, I22.0, I22.1, I22.8, I22.9;<br>I23.X, I23.1, I23.2, I23.3, I23.6, I23.8;<br>I24.1;<br>I25.2 | Acute myocardial infarction; Subsequent myocardial infarction; Certain current complications following acute myocardial infarction; Dressler's syndrome; Old myocardial infarction |
| Diagnoses – ICD 10 | 41270 | I21.X, I21.0, I21.1, I21.2, I21.3, I21.4, I21.9;<br>I22.X, I22.0, I22.1, I22.8, I22.9;<br>I23.X, I23.1, I23.2, I23.3, I23.6, I23.8;<br>I24.1;<br>I25.2 | Acute myocardial infarction; Subsequent myocardial infarction; Certain current complications following acute myocardial infarction; Dressler's syndrome; Old myocardial infarction |
| Operative procedures – OPCS4 | 41272 | K40.1, K40.2, K40.3, K40.4;<br>K41.1, K41.2, K41.3, K41.4;<br>K45.1, K45.2, K45.3, K45.4, K45.5;<br>K49.1, K49.2, K49.8, K49.9;<br>K75.1, K75.2, K75.3, K75.4, K75.8, K75.9;<br>K50.2; | Saphenous vein graft replacement of (one – four or more) coronary artery; Other autograft replacement of (one – four or more) coronary artery; Connection of mammary/thoracic artery to the coronary artery; Percutaneous transluminal balloon angioplasty ± insertion of stent into the coronary artery; Percutaneous transluminal coronary thrombolysis using streptokinase |
| ***Heart Failure*** | | | |
| Non-cancer Illness Codes | 20002 | 1076, 1079 | Heart failure/pulmonary oedema, Cardiomyopathy |
| Underlying (primary) cause of death: ICD10 | 40001 | I11.0;<br>I13.0, I13.2;<br>I25.5;<br>I42.0, I42.5, I42.8, I42.9;<br>I50.0, I50.1, I50.9 | Hypertensive heart disease with (congestive) heart failure; Hypertensive heart and renal disease with (congestive) heart failure and/or renal failure; Ischaemic cardiomyopathy; Dilated cardiomyopathy, Other restrictive/unspecified cardiomyopathies; Heart failure |
| Contributory (secondary) causes of death: ICD10 | 40002 | I11.0;<br>I13.0, I13.2;<br>I25.5;<br>I42.0, I42.5, I42.8, I42.9;<br>I50.0, I50.1, I50.9 | Hypertensive heart disease with (congestive) heart failure; Hypertensive heart and renal disease with (congestive) heart failure and/or renal failure; Ischaemic cardiomyopathy; Dilated cardiomyopathy, Other restrictive/unspecified cardiomyopathy; Heart failure |
| Diagnoses – ICD 10 | 41270 | I11.0;<br>I13.0, I13.2;<br>I25.5;<br>I42.0, I42.5, I42.8, I42.9;<br>I50.0, I50.1, I50.9 | Hypertensive heart disease with (congestive) heart failure; Hypertensive heart and renal disease with (congestive) heart failure and/or renal failure; Ischaemic cardiomyopathy; Dilated cardiomyopathy, Other restrictive/unspecified cardiomyopathy; Heart failure |
| Diagnoses – ICD 9 | 41271 | 4254; 4280, 4281, 4289 | Other primary cardiomyopathies; Heart failure |
| ***Hypertension*** | | | |
| Non-cancer Illness Codes | 20002 | 1065, 1072 | Hypertension, Essential hypertension |
| Underlying (primary) cause of death: ICD10 | 40001 | I10;<br>I11.X, I11.0, I11.9;<br>I12.X, I12.0, I12.9;<br>I13.X, I1.30, I13.1, I13.2, I13.9;<br>I15.0, I15.1, I15.2, I15.9 | Essential (primary) hypertension; Hypertensive heart disease; Hypertensive renal disease; Hypertensive heart and renal disease; Secondary hypertension |
| Contributory (secondary) causes of death: ICD10 | 40002 | I10;<br>I11.X, I11.0, I11.9;<br>I12.X, I12.0, I12.9;<br>I13.X, I1.30, I13.1, I13.2, I13.9;<br>I15.0, I15.1, I15.2, I15.9 | Essential (primary) hypertension; Hypertensive heart disease; Hypertensive renal disease; Hypertensive heart and renal disease; Secondary hypertension |
| Diagnoses – ICD 10 | 41270 | I10;<br>I11.X, I11.0, I11.9;<br>I12.X, I12.0, I12.9;<br>I13.X, I1.30, I13.1, I13.2, I13.9;<br>I15.0, I15.1, I15.2, I15.9 | Essential (primary) hypertension; Hypertensive heart disease; Hypertensive renal disease; Hypertensive heart and renal disease; Secondary hypertension |

**Supplementary Table 1: Definitions of incident endpoints and relevant UKB data fields and codes.** Endpoint definitions were adapted as published elsewhere[1, 2].

| Field names | Data fields | Data coding | Meaning |
|---|---|---|---|
| ***Sociodemographics*** | | | |
| ***Age*** | | | |
| Age at recruitment | 21022 | Integer | Years |
| ***Sex*** | | | |
| Sex | 31 | 0; 1 | Female; Male |
| ***Ethnicity*** | | | |
| Ethnic background | 21000 | 1, 1001, 1002, 1003;<br>4, 4001, 4002, 4003, 4004;<br>2, 2001, 2002, 2003, 2004;<br>3, 3001, 3002, 3003, 3004;<br>5; 6 | White;<br>Black or Black British;<br>Other (Mixed; Asian or Asian British; Chinese; Other ethnic group) |
| ***Higher education*** | | | |
| Qualifications | 6138 | 1 | College or University degree |
| ***Lifestyle factors*** | | | |
| ***Smoking status*** | | | |
| Smoking status | 20116 | 0; 1; 2 | Never; Previous; Current |
| ***Current smoker*** | | | |
| Smoking status | 20116 | 2 | Current |
| ***Alcohol consumption*** | | | |
| Alcohol drinker status | 20117 | 0; 1; 2 | Never; Previous; Current |
| ***Daily physical activity*** | | | |
| Summed minutes activity | 22034 | Integer | Minutes |
| ***Clinical chemistry*** | | | |
| Cholesterol | 30690 | Continuous | mmol/L |
| HDL cholesterol | 30760 | Continuous | mmol/L |
| LDL cholesterol | 30780 | Continuous | mmol/L |
| Triglycerides | 30870 | Continuous | mmol/L |
| Glucose | 30740 | Continuous | mmol/L |
| Glycated haemoglobin/HbA1c | 30750 | Continuous | mmol/mol |
| Creatinine | 30700 | Continuous | µmol/L |
| C-reactive protein | 30710 | Continuous | mg/L |
| ***Physical measurements*** | | | |
| ***Systolic blood pressure (SBP)*** | | | |
| Systolic blood pressure, automated reading | 4080 | Integer | mmHg |
| Systolic blood pressure, manual reading | 93 | Integer | mmHg |
| ***Body mass index (BMI)*** | | | |
| Body mass index | 21001 | Continuous | kg/m$^2$ |
| ***Waist/hip ratio*** | | | |
| Waist circumference | 48 | Continuous | cm |
| Hip circumference | 49 | Continuous | cm |
| ***Medication*** | | | |
| ***Lipid lowering medication*** | | | |
| Medication for cholesterol, blood pressure, diabetes, or take exogenous hormones (Sexed: Female only) | 6153 | 1 | Cholesterol lowering medication |

| | | | |
|---|---|---|---|
| Medication for cholesterol, blood pressure or diabetes (Sexed: Male only) | 6177 | 1 | Cholesterol lowering medication |
| Treatment/medication code | 20003 | 1141146234, 1140888594, 1140888648, 1141192410, 1140861958, 1140865576, 1140865576, 1140861936, 1140861944, 1140862028, 1141175908, 1140851880, 1140851882, 1140861324, 1140861868, 1140861892, 1141162544, 1141192736, 1141192740, 1141157416, 1140861924, 1141157260, 1140861926, 1140861928, 1140861922, 1140861942, 1140861946, 1140861954, 1140862026, 1141168568, 1141171548, 1141201306, 1140888590, 1140861848, 1140861856, 1141157262, 1140861858, 1140926582, 1140861866, 1141188546, 1140861876, 1140861878, 1140861884, 1141181868, 1141172214, 1141182910, 1140865752, 1141157494, 1141145830 | Lipid modifying medication |
| ***Treatment for diabetes/DM medication*** | | | |
| Medication for cholesterol, blood pressure, diabetes, or take exogenous hormones (Sexed: Female only) | 6153 | 3 | Insulin |
| Medication for cholesterol, blood pressure or diabetes (Sexed: Male only) | 6177 | 3 | Insulin |
| Treatment/medication code | 20003 | 1140857494, 1140857496, 1140857500, 1140857502, 1140857506, 1140857584, 1140857586, 1140857590, 1140868902, 1140868908, 1140874646, 1140874650, 1140874652, 1140874658, 1140874660, 1140874664, 1140874666, 1140874674, 1140874678, 1140874680, 1140874686, 1140874690, 1140874706, 1140874712, 1140874716, 1140874718, 1140874724, 1140874726, 1140874728, 1140874732, 1140874736, 1140874740, 1140874744, 1140874746, 1140882964, 1140883066, 1140884600, 1140910564, 1140910566, 1140910818, 1140921964, 1141152590, 1141153254, 1141153262, 1141156984, 1141157284, 1141168660, 1141168668, 1141169504, 1141171508, 1141171646, 1141171652, 1141173786, 1141173882, 1141177600, 1141177606, 1141189090, 1141189094 | Diabetes mellitus medication |
| ***Treatment for hypertension/HT medication*** | | | |
| Medication for cholesterol, blood pressure, diabetes, or take exogenous hormones (Sexed: Female only) | 6153 | 2 | Blood pressure medication |
| Medication for cholesterol, blood pressure or diabetes (Sexed: Male only) | 6177 | 2 | Blood pressure medication |

| | | | |
|---|---|---|---|
| Treatment/medication code | 20003 | 1140860332, 1140860334, 1140860336, 1140860338, 1140860340, 1140860342, 1140860348, 1140860352, 1140860356, 1140860358, 1140860362, 1140860380, 1140860390, 1140860394, 1140860396, 1140860398, 1140860402, 1140860410, 1140860418, 1140860422, 1140860426, 1140860434, 1140860478, 1140860492, 1140860498, 1140860520, 1140860532, 1140860552, 1140860558, 1140860562, 1140860564, 1140860580, 1140860628, 1140860632, 1140860638, 1140860654, 1140860658, 1140860706, 1140860714, 1140860728, 1140860736, 1140860738, 1140860758, 1140860764, 1140860776, 1140860784, 1140860790, 1140860828, 1140860830, 1140860834, 1140860836, 1140860838, 1140860846, 1140860848, 1140860862, 1140860878, 1140860882, 1140860912, 1140860918, 1140860938, 1140860942, 1140860952, 1140860972, 1140860976, 1140860982, 1140860988, 1140860994, 1140861008, 1140861010, 1140861016, 1140861022, 1140861024, 1140861068, 1140861070, 1140861088, 1140861090, 1140861106, 1140861120, 1140861128, 1140861130, 1140861136, 1140861138, 1140861190, 1140861194, 1140861202, 1140861266, 1140861268, 1140861326, 1140861384, 1140864950, 1140864952, 1140866072, 1140866084, 1140866086, 1140866090, 1140866092, 1140866094, 1140866104, 1140866108, 1140866110, 1140866116, 1140866122, 1140866136, 1140866138, 1140866140, 1140866144, 1140866146, 1140866162, 1140866164, 1140866168, 1140866182, 1140866192, 1140866202, 1140866206, 1140866210, 1140866212, 1140866220, 1140866230, 1140866232, 1140866236, 1140866244, 1140866248, 1140866282, 1140866306, 1140866308, 1140866312, 1140866318, 1140866330, 1140866332, 1140866334, 1140866340, 1140866352, 1140866360, 1140866388, 1140866390, 1140866396, 1140866400, 1140866406, 1140866408, 1140866410, 1140866412, 1140866416, 1140866422, 1140866426, 1140866438, 1140866440, 1140866442, 1140866448, 1140866450, 1140866460, 1140866466, 1140866484, 1140866554, 1140866692, 1140866704, 1140866712, 1140866724, 1140866756, 1140866758, 1140866764, 1140866766, 1140866778, 1140866798, 1140866800, 1140866802, 1140866804, 1140875808, 1140879762, 1140879778, 1140879782, 1140879786, 1140879794, 1140879806, 1140879810, 1140879818, 1140879822, 1140879824, 1140879834, 1140879842, 1140879854, 1140879866, 1140888510, 1140888556, 1140888560, 1140888578, 1140888582, 1140888586, 1140888760, 1140888762, 1140909368, 1140911698, 1140916356, 1140923572, 1140923712, 1140923718, 1140926778, 1140926780, 1141145668, 1141151016, 1141151018, 1141151382, 1141152600, 1141153026, 1141153032, 1141153328, 1141156754, 1141156808, 1141157252, 1141157254, 1141164148, 1141164154, 1141164276, 1141165476, 1141166006, 1141167822, 1141167832, 1141171152, 1141172682, 1141172686, 1141172698, 1141173888, 1141180592, 1141187790, 1141190160, 1141192064, 1141193282, 1141193346, 1141194804, 1141194808, 1141194810, 1141201038, 1141201040, 1140860382, 1140860386, 1140860404, 1140860406, 1140860454, 1140860470, 1140860534, 1140860544, 1140860590, 1140860610, 1140860690, 1140860696, 1140860750, 1140860752, 1140860802, 1140860806, 1140860840, 1140860842, 1140860892, 1140860904, 1140860954, 1140860966, 1140861000, 1140861002, 1140861034, 1140861046, 1140861110, 1140861114, 1140861166, 1140861176, 1140861276, 1140861282, 1140866074, 1140866078, 1140866096, 1140866102, 1140866128, 1140866132, 1140866156, 1140866158, 1140866194, 1140866200, 1140866222, 1140866226, 1140866262, 1140866280, 1140866324, 1140866328, 1140866354, 1140866356, 1140866402, 1140866404, 1140866418, 1140866420, 1140866444, 1140866446, 1140866506, 1140866546, 1140866726, 1140866738, 1140866782, 1140866784, 1140879758, 1140879760, 1140879798, 1140879802, 1140879826, 1140879830, 1140888512, 1140888552, 1140888646, 1140888686, 1140916362, 1140917428, 1141145658, 1141145660, 1141152998, 1141153006, 1141156836, 1141156846, 1141164280, 1141165470, 1141171336, 1141171344, 1141180598, 1141187788, 1141194794, 1141194800 | Blood pressure medication |

***Family history of heart disease***

| | | | |
|---|---|---|---|
| Illnesses of (biological) father | 20107 | 1 | Heart disease |
| Illnesses of (biological) mother | 20110 | 1 | Heart disease |
| ***1H-NMR Metabolites*** | | | |
| NMR metabolic biomarker measurements | 20280, 23400-23440, 23442-23450, 23460-23578 | Continuous | |

**Supplementary Table 2: Definitions of baseline characteristic and predictor variables and relevant UKB data fields and codes.** Medication codes are adapted from definitions used elsewhere[3, 4].

| Variable | HR | Upper 95% CI | Lower 95% CI | P-value | Adjusted P-value |
|---|---|---|---|---|---|
| ***Glycolysis related metabolites*** | | | | | |
| Citrate | 0.988371214 | 0.936680219 | 1.042914792 | 0.66953317 | 0.690070997 |
| Glucose | 1.097840348 | 1.039778425 | 1.159144488 | 0.000759956 | 0.001798205 |
| Lactate | 1.026441176 | 0.973844821 | 1.081878205 | 0.330839483 | 0.378102267 |
| Pyruvate | 1.066646857 | 1.010204036 | 1.126243291 | 0.020020822 | 0.032033316 |
| ***Amino acids*** | | | | | |
| Alanine | 0.941718182 | 0.894074359 | 0.99190087 | 0.023392329 | 0.036054232 |
| Glutamine | 0.886937052 | 0.841925895 | 0.934354602 | 6.33E-06 | 3.00E-05 |
| Glycine | 0.86748864 | 0.818393268 | 0.919529241 | 1.73E-06 | 1.10E-05 |
| Histidine | 0.854689112 | 0.811332651 | 0.900362482 | 3.39E-09 | 6.33E-08 |
| Isoleucine | 1.037767357 | 0.983959705 | 1.094517472 | 0.172348809 | 0.205351773 |
| Leucine | 1.017008809 | 0.963067748 | 1.073971088 | 0.544137626 | 0.578576716 |
| Phenylalanine | 1.095324929 | 1.039542566 | 1.154100601 | 0.000639894 | 0.001558002 |
| Total Concentration of Branched-Chain Amino Acids (Leucine + Isoleucine + Valine) | 1.026732385 | 0.972760867 | 1.083698396 | 0.338286469 | 0.38142367 |
| Tyrosine | 1.112551144 | 1.055939018 | 1.17219842 | 6.26E-05 | 0.000202335 |
| Valine | 1.024499701 | 0.970559225 | 1.081438011 | 0.380434136 | 0.4177316 |
| ***Apolipoproteins*** | | | | | |
| Apolipoprotein A1 | 0.858078116 | 0.809865789 | 0.909160582 | 2.13E-07 | 1.70E-06 |
| Apolipoprotein B | 0.974572097 | 0.922529646 | 1.029550408 | 0.357632851 | 0.400548793 |
| **Lipoprotein particle sizes** | | | | | |
| Average Diameter for HDL Particles | 0.891180174 | 0.838298056 | 0.94739824 | 0.00022315 | 0.000595067 |
| Average Diameter for LDL Particles | 0.929528916 | 0.883047219 | 0.978457309 | 0.005237856 | 0.009777331 |
| Average Diameter for VLDL Particles | 1.028413064 | 0.97138857 | 1.088785128 | 0.335748254 | 0.38111964 |
| ***Other lipids*** | | | | | |
| Phosphatidylcholines | 0.894776734 | 0.845550726 | 0.946868567 | 0.000117654 | 0.000335015 |
| Phosphoglycerides | 0.922235518 | 0.871789938 | 0.975600101 | 0.004792486 | 0.009149292 |
| Sphingomyelins | 0.925711961 | 0.87509037 | 0.979261873 | 0.007137941 | 0.012798832 |
| Total Cholines | 0.905733316 | 0.856076321 | 0.958270682 | 0.000578253 | 0.001449947 |
| ***Fatty acids*** | | | | | |
| Degree of Unsaturation | 0.801352287 | 0.760539739 | 0.844354942 | 1.01E-16 | 8.49E-15 |
| Docosahexaenoic Acid | 0.805534512 | 0.764578858 | 0.848684008 | 4.57E-16 | 1.92E-14 |
| Linoleic Acid | 0.903502076 | 0.856479022 | 0.953106825 | 0.000198306 | 0.000546153 |
| Monounsaturated Fatty Acids | 1.112461776 | 1.054857459 | 1.173211786 | 8.54E-05 | 0.000261107 |
| Omega-3 Fatty Acids | 0.845794127 | 0.802530482 | 0.891390073 | 4.06E-10 | 9.74E-09 |
| Omega-6 Fatty Acids | 0.927974951 | 0.878930597 | 0.979755982 | 0.006971811 | 0.012731133 |
| Polyunsaturated Fatty Acids | 0.898021723 | 0.850240432 | 0.948488198 | 0.000115356 | 0.000335015 |
| Saturated Fatty Acids | 1.057445359 | 1.002478969 | 1.115425583 | 0.040279411 | 0.057258952 |
| Total Fatty Acids | 1.024603046 | 0.97069151 | 1.081508791 | 0.378139776 | 0.4177316 |
| ***Lipoprotein particle concentrations*** | | | | | |
| Concentration of HDL Particles | 0.846279683 | 0.800365675 | 0.894827607 | 4.50E-09 | 6.88E-08 |
| Concentration of LDL Particles | 0.959023492 | 0.90797164 | 1.012945798 | 0.133848097 | 0.162945509 |
| Concentration of VLDL Particles | 1.05194853 | 0.994626552 | 1.112574069 | 0.076477964 | 0.1001797 |
| Total Concentration of Lipoprotein Particles | 0.850953469 | 0.804921079 | 0.899618392 | 1.28E-08 | 1.80E-07 |
| ***Lipoprotein particles*** | | | | | |
| Concentration of Chylomicrons and Extremely Large VLDL Particles | 1.156852459 | 1.085768419 | 1.232590291 | 6.69E-06 | 3.00E-05 |
| Concentration of IDL Particles | 0.990565567 | 0.937906203 | 1.046181526 | 0.733773124 | 0.747114454 |
| Concentration of Large HDL Particles | 0.851316498 | 0.802424493 | 0.903187509 | 9.60E-08 | 9.48E-07 |
| Concentration of Large LDL Particles | 0.952301077 | 0.902100353 | 1.005295406 | 0.076923698 | 0.1001797 |
| Concentration of Large VLDL Particles | 1.053465974 | 0.993844027 | 1.116664716 | 0.079734436 | 0.101546827 |
| Concentration of Medium HDL Particles | 0.860459607 | 0.813083915 | 0.910595722 | 1.98E-07 | 1.66E-06 |
| Concentration of Medium LDL Particles | 0.965506288 | 0.913616558 | 1.020343145 | 0.212971415 | 0.248466651 |
| Concentration of Medium VLDL Particles | 0.984951194 | 0.931118402 | 1.04189634 | 0.596971879 | 0.63076274 |
| Concentration of Small HDL Particles | 0.898610491 | 0.851937656 | 0.947840266 | 8.55E-05 | 0.000261107 |
| Concentration of Small LDL Particles | 0.987759995 | 0.934899246 | 1.043609578 | 0.660759484 | 0.685232057 |
| Concentration of Small VLDL Particles | 1.049553233 | 0.992191892 | 1.110230791 | 0.091679271 | 0.114941175 |
| Concentration of Very Large HDL Particles | 0.893696385 | 0.841548512 | 0.949075684 | 0.000248466 | 0.000652224 |
| Concentration of Very Large VLDL Particles | 1.09427242 | 1.02804951 | 1.164761149 | 0.004676767 | 0.009030999 |
| Concentration of Very Small VLDL Particles | 1.082975142 | 1.024187666 | 1.145136966 | 0.005122069 | 0.009668624 |
| ***Cholesterol*** | | | | | |
| Clinical LDL Cholesterol | 0.913112325 | 0.86496976 | 0.963934415 | 0.001004975 | 0.002251143 |

| | | | | | |
|---|---|---|---|---|---|
| HDL Cholesterol | 0.82553336 | 0.778388951 | 0.875533147 | 1.65E-10 | 4.63E-09 |
| LDL Cholesterol | 0.907382023 | 0.85958502 | 0.957836767 | 0.000431197 | 0.001114478 |
| Remnant Cholesterol (Non-HDL, Non-LDL -Cholesterol) | 0.977950903 | 0.925402171 | 1.033483601 | 0.428825324 | 0.464791319 |
| Total Cholesterol | 0.893231495 | 0.845869347 | 0.943245557 | 4.87E-05 | 0.000163513 |
| Total Cholesterol Minus HDL-C | 0.939548143 | 0.889589691 | 0.992312212 | 0.025299368 | 0.038290935 |
| VLDL Cholesterol | 1.042372368 | 0.98492693 | 1.103168286 | 0.15133195 | 0.18159834 |
| Cholesterol in Chylomicrons and Extremely Large VLDL | 1.151319317 | 1.083840763 | 1.222999002 | 4.82E-06 | 2.53E-05 |
| Cholesterol in IDL | 0.908705255 | 0.860552724 | 0.959552176 | 0.000568349 | 0.001446706 |
| Cholesterol in Large HDL | 0.849563241 | 0.800762945 | 0.901337538 | 6.61E-08 | 7.40E-07 |
| Cholesterol in Large LDL | 0.892269995 | 0.844599575 | 0.942631002 | 4.72E-05 | 0.000161927 |
| Cholesterol in Large VLDL | 1.054281375 | 0.992512439 | 1.119894496 | 0.086165726 | 0.108840917 |
| Cholesterol in Medium HDL | 0.842130985 | 0.795230633 | 0.89179738 | 4.18E-09 | 6.88E-08 |
| Cholesterol in Medium LDL | 0.932959476 | 0.883228696 | 0.985490381 | 0.013030776 | 0.021891703 |
| Cholesterol in Medium VLDL | 0.950057417 | 0.898506388 | 1.004566142 | 0.071874612 | 0.096599478 |
| Cholesterol in Small HDL | 0.883312637 | 0.83747147 | 0.931663038 | 5.04E-06 | 2.56E-05 |
| Cholesterol in Small LDL | 0.937988493 | 0.888193557 | 0.990575089 | 0.021435962 | 0.033973978 |
| Cholesterol in Small VLDL | 1.036852524 | 0.979586714 | 1.097466045 | 0.211861131 | 0.248466651 |
| Cholesterol in Very Large HDL | 0.885677384 | 0.834329332 | 0.940185606 | 6.78E-05 | 0.000214772 |
| Cholesterol in Very Large VLDL | 1.08667743 | 1.021510812 | 1.15600131 | 0.008426673 | 0.014746679 |
| Cholesterol in Very Small VLDL | 1.02531002 | 0.969212497 | 1.08465444 | 0.383935679 | 0.418838923 |
| ***Cholesteryl esters*** | | | | | |
| Cholesteryl Esters in HDL | 0.821483566 | 0.775050906 | 0.870697967 | 3.49E-11 | 1.17E-09 |
| Cholesteryl Esters in LDL | 0.91566169 | 0.867265394 | 0.966758661 | 0.001471908 | 0.003170264 |
| Cholesteryl Esters in VLDL | 1.033335888 | 0.976603099 | 1.093364395 | 0.255031843 | 0.29548517 |
| Total Esterified Cholesterol | 0.883417713 | 0.836648399 | 0.932801468 | 7.95E-06 | 3.18E-05 |
| Cholesteryl Esters in Chylomicrons and Extremely Large VLDL | 1.149902766 | 1.081583405 | 1.222537592 | 7.84E-06 | 3.18E-05 |
| Cholesteryl Esters in IDL | 0.902446408 | 0.854740078 | 0.952815412 | 0.000212054 | 0.000574598 |
| Cholesteryl Esters in Large HDL | 0.845582897 | 0.797345892 | 0.896738094 | 2.18E-08 | 2.82E-07 |
| Cholesteryl Esters in Large LDL | 0.89606738 | 0.848044351 | 0.946809856 | 9.43E-05 | 0.000282932 |
| Cholesteryl Esters in Large VLDL | 1.0403888 | 0.979360916 | 1.105219574 | 0.199219707 | 0.235696555 |
| Cholesteryl Esters in Medium HDL | 0.840426287 | 0.793650997 | 0.889958365 | 2.68E-09 | 5.63E-08 |
| Cholesteryl Esters in Medium LDL | 0.948826912 | 0.897841314 | 1.002707823 | 0.062320892 | 0.085121218 |
| Cholesteryl Esters in Medium VLDL | 0.929473618 | 0.877038443 | 0.985043714 | 0.013564318 | 0.02256243 |
| Cholesteryl Esters in Small HDL | 0.871187259 | 0.82328002 | 0.921882254 | 1.77E-06 | 1.10E-05 |
| Cholesteryl Esters in Small LDL | 0.958766636 | 0.907590053 | 1.01282893 | 0.132452125 | 0.162423044 |
| Cholesteryl Esters in Small VLDL | 1.064994577 | 1.005479402 | 1.128032505 | 0.031856244 | 0.046946043 |
| Cholesteryl Esters in Very Large HDL | 0.865981083 | 0.813152646 | 0.922241649 | 7.45E-06 | 3.13E-05 |
| Cholesteryl Esters in Very Large VLDL | 1.075356882 | 1.010213581 | 1.144700927 | 0.0226862 | 0.035619455 |
| Cholesteryl Esters in Very Small VLDL | 1.002059553 | 0.947317274 | 1.059965203 | 0.942777067 | 0.948422438 |
| ***Free cholesterol*** | | | | | |
| Free Cholesterol in HDL | 0.85060013 | 0.801086491 | 0.903174113 | 1.24E-07 | 1.15E-06 |
| Free Cholesterol in LDL | 0.887552841 | 0.840787642 | 0.936919153 | 1.57E-05 | 5.84E-05 |
| Free Cholesterol in VLDL | 1.05212671 | 0.993984104 | 1.11367034 | 0.079786793 | 0.101546827 |
| Total Free Cholesterol | 0.920197978 | 0.871213396 | 0.971936753 | 0.002883932 | 0.005981488 |
| Free Cholesterol in Chylomicrons and Extremely Large VLDL | 1.148770712 | 1.081568302 | 1.220148693 | 6.50E-06 | 3.00E-05 |
| Free Cholesterol in IDL | 0.928803786 | 0.879406491 | 0.980975785 | 0.0080774 | 0.014284245 |
| Free Cholesterol in Large HDL | 0.864704699 | 0.81268036 | 0.920059414 | 4.40E-06 | 2.38E-05 |
| Free Cholesterol in Large LDL | 0.884518264 | 0.837745878 | 0.933902011 | 9.56E-06 | 3.65E-05 |
| Free Cholesterol in Large VLDL | 1.066285217 | 1.003785024 | 1.132676953 | 0.03729234 | 0.054009595 |
| Free Cholesterol in Medium HDL | 0.85276373 | 0.805197361 | 0.903140044 | 5.36E-08 | 6.43E-07 |
| Free Cholesterol in Medium LDL | 0.894972211 | 0.847654749 | 0.94493101 | 6.23E-05 | 0.000202335 |
| Free Cholesterol in Medium VLDL | 0.974618825 | 0.921128869 | 1.031214943 | 0.372031134 | 0.413915433 |
| Free Cholesterol in Small HDL | 0.910735094 | 0.862520159 | 0.961645247 | 0.000753858 | 0.001798205 |
| Free Cholesterol in Small LDL | 0.892213719 | 0.844827542 | 0.942257775 | 4.20E-05 | 0.000147107 |
| Free Cholesterol in Small VLDL | 0.988892899 | 0.935135421 | 1.045740696 | 0.695314633 | 0.712273527 |
| Free Cholesterol in Very Large HDL | 0.944972934 | 0.890939109 | 1.002283813 | 0.0595611 | 0.082018563 |
| Free Cholesterol in Very Large VLDL | 1.096805548 | 1.031047357 | 1.166757667 | 0.003398103 | 0.006878088 |
| Free Cholesterol in Very Small VLDL | 1.078100908 | 1.01923214 | 1.140369815 | 0.008667788 | 0.015012252 |
| ***Phospholipids*** | | | | | |
| Phospholipids in HDL | 0.874197506 | 0.824476388 | 0.92691712 | 6.79E-06 | 3.00E-05 |

| | | | | | |
|---|---|---|---|---|---|
| Phospholipids in LDL | 0.928379233 | 0.879433992 | 0.98004854 | 0.007161251 | 0.012798832 |
| Phospholipids in VLDL | 1.06817764 | 1.009139059 | 1.130670208 | 0.022991504 | 0.035764562 |
| Total Phospholipids in Lipoprotein Particles | 0.921362547 | 0.871293857 | 0.974308423 | 0.004066528 | 0.008133056 |
| Phospholipids in Chylomicrons and Extremely Large VLDL | 1.178874027 | 1.097633141 | 1.266127924 | 6.27E-06 | 3.00E-05 |
| Phospholipids in IDL | 0.954469776 | 0.90289439 | 1.008991266 | 0.100146026 | 0.124626166 |
| Phospholipids in Large HDL | 0.864771269 | 0.814368247 | 0.918293843 | 2.12E-06 | 1.27E-05 |
| Phospholipids in Large LDL | 0.92080712 | 0.872334724 | 0.971972946 | 0.002787267 | 0.005853261 |
| Phospholipids in Large VLDL | 1.094667738 | 1.014365239 | 1.181327406 | 0.019971305 | 0.032033316 |
| Phospholipids in Medium HDL | 0.894047701 | 0.84453058 | 0.946468145 | 0.000116918 | 0.000335015 |
| Phospholipids in Medium LDL | 0.939316081 | 0.889453579 | 0.99197386 | 0.024478661 | 0.037385591 |
| Phospholipids in Medium VLDL | 0.986469424 | 0.932099392 | 1.044010899 | 0.637664124 | 0.665388651 |
| Phospholipids in Small HDL | 0.945328404 | 0.895808559 | 0.997585681 | 0.040558424 | 0.057258952 |
| Phospholipids in Small LDL | 0.942342067 | 0.892794353 | 0.994639549 | 0.031161468 | 0.046328554 |
| Phospholipids in Small VLDL | 1.014017095 | 0.958392581 | 1.072870021 | 0.628686059 | 0.660120362 |
| Phospholipids in Very Large HDL | 0.909778685 | 0.850777134 | 0.972872005 | 0.005711619 | 0.010544527 |
| Phospholipids in Very Large VLDL | 1.12070328 | 1.040914734 | 1.206607804 | 0.002493638 | 0.005302926 |
| Phospholipids in Very Small VLDL | 1.114779632 | 1.054233079 | 1.178803484 | 0.00013696 | 0.000383487 |
| ***Total lipids*** | | | | | |
| Total Lipids in HDL | 0.855152116 | 0.806230034 | 0.907042794 | 1.93E-07 | 1.66E-06 |
| Total Lipids in LDL | 0.924409725 | 0.875586721 | 0.975955115 | 0.004524107 | 0.008941763 |
| Total Lipids in Lipoprotein Particles | 0.942684941 | 0.892607711 | 0.995571613 | 0.034062943 | 0.049761517 |
| Total Lipids in VLDL | 1.057340002 | 0.999543792 | 1.118478137 | 0.051888968 | 0.072044186 |
| Total Lipids in Chylomicrons and Extremely Large VLDL | 1.129975487 | 1.06592952 | 1.197869631 | 4.05E-05 | 0.000144756 |
| Total Lipids in IDL | 0.935818928 | 0.885598516 | 0.988887233 | 0.018420802 | 0.03004558 |
| Total Lipids in Large HDL | 0.858557159 | 0.809168886 | 0.910959884 | 4.53E-07 | 3.31E-06 |
| Total Lipids in Large LDL | 0.912510289 | 0.864385725 | 0.963314182 | 0.000926235 | 0.00213161 |
| Total Lipids in Large VLDL | 1.0318546 | 0.973896332 | 1.093262065 | 0.28770356 | 0.331056152 |
| Total Lipids in Medium HDL | 0.876130261 | 0.828177703 | 0.926859335 | 4.13E-06 | 2.31E-05 |
| Total Lipids in Medium LDL | 0.944238539 | 0.893852547 | 0.997464762 | 0.040297055 | 0.057258952 |
| Total Lipids in Medium VLDL | 0.980392822 | 0.926985334 | 1.036877337 | 0.488395744 | 0.525964648 |
| Total Lipids in Small HDL | 0.934941069 | 0.886106403 | 0.986467088 | 0.01398056 | 0.023026804 |
| Total Lipids in Small LDL | 0.949902168 | 0.899490441 | 1.003139208 | 0.064702254 | 0.087661119 |
| Total Lipids in Small VLDL | 1.042752561 | 0.985521031 | 1.103307662 | 0.14606759 | 0.176542123 |
| Total Lipids in Very Large HDL | 0.904847204 | 0.852304304 | 0.960629272 | 0.001053085 | 0.002327872 |
| Total Lipids in Very Large VLDL | 1.081016059 | 1.018099031 | 1.147821267 | 0.010888675 | 0.0186663 |
| Total Lipids in Very Small VLDL | 1.084421699 | 1.025258986 | 1.146998405 | 0.004633739 | 0.009030999 |
| ***Triglycerides*** | | | | | |
| Total Triglycerides | 1.07270746 | 1.01553533 | 1.13309824 | 0.012017562 | 0.020393439 |
| Triglycerides in HDL | 1.096161454 | 1.03789655 | 1.157697203 | 0.000985144 | 0.002236542 |
| Triglycerides in LDL | 1.134785231 | 1.076413091 | 1.196322798 | 2.69E-06 | 1.56E-05 |
| Triglycerides in VLDL | 1.052001003 | 0.995117635 | 1.112135963 | 0.073873273 | 0.098497697 |
| Triglycerides in Chylomicrons and Extremely Large VLDL | 1.122295066 | 1.046709483 | 1.203338878 | 0.001181812 | 0.002578499 |
| Triglycerides in IDL | 1.156986971 | 1.096908136 | 1.220356389 | 8.34E-08 | 8.75E-07 |
| Triglycerides in Large HDL | 1.019778644 | 0.963969974 | 1.078818336 | 0.495198551 | 0.529893991 |
| Triglycerides in Large LDL | 1.14608936 | 1.087034847 | 1.208352082 | 4.38E-07 | 3.31E-06 |
| Triglycerides in Large VLDL | 0.998915904 | 0.944082285 | 1.056934337 | 0.969962154 | 0.969962154 |
| Triglycerides in Medium HDL | 1.098682746 | 1.039439091 | 1.16130304 | 0.000875705 | 0.002043312 |
| Triglycerides in Medium LDL | 1.118449583 | 1.060784716 | 1.179249145 | 3.40E-05 | 0.000124192 |
| Triglycerides in Medium VLDL | 0.993183958 | 0.939496726 | 1.04993913 | 0.809385182 | 0.819136811 |
| Triglycerides in Small HDL | 1.139727128 | 1.076512537 | 1.206653785 | 7.05E-06 | 3.03E-05 |
| Triglycerides in Small LDL | 1.097514646 | 1.040639872 | 1.157497834 | 0.000609776 | 0.001506504 |
| Triglycerides in Small VLDL | 1.050902208 | 0.994202307 | 1.110835735 | 0.079346613 | 0.101546827 |
| Triglycerides in Very Large HDL | 1.0508705 | 0.994805122 | 1.110095619 | 0.076100604 | 0.1001797 |
| Triglycerides in Very Large VLDL | 1.067398257 | 1.00602385 | 1.132516926 | 0.030870449 | 0.046305674 |
| Triglycerides in Very Small VLDL | 1.142584189 | 1.081993883 | 1.206567476 | 1.63E-06 | 1.09E-05 |
| ***Ketone bodies*** | | | | | |
| 3-Hydroxybutyrate | 1.131379604 | 1.071573831 | 1.194523206 | 8.40E-06 | 3.28E-05 |
| Acetate | 0.946563831 | 0.89673331 | 0.999163381 | 0.04655818 | 0.065181452 |
| Acetoacetate | 1.150158719 | 1.088365986 | 1.215459776 | 6.86E-07 | 4.80E-06 |
| Acetone | 1.07824243 | 1.025465487 | 1.133735607 | 0.003260504 | 0.006680058 |
| ***Fluid balance*** | | | | | |

| | | | | | |
|---|---|---|---|---|---|
| Albumin | 0.800776408 | 0.760166036 | 0.843556309 | 5.92E-17 | 8.49E-15 |
| Creatinine | 1.051356491 | 0.988916955 | 1.117738416 | 0.10889179 | 0.134513388 |
| ***Inflammation*** | | | | | |
| Glycoprotein Acetyls | 1.250087272 | 1.185536122 | 1.318153161 | 1.56E-16 | 8.75E-15 |

**Supplementary Table 3: Results of per-metabolite COX-regressions after adjustment for age and sex.** P-values were adjusted following the Benjamini-Hochberg method. HR = Hazard ratio. CI = Confidence interval.

| Variable | HR | Upper 95% CI | Lower 95% CI | p-value | Adjusted p-value |
|---|---|---|---|---|---|
| ***Glycolysis related metabolites*** | | | | | |
| Citrate | 1.002404737 | 0.949075018 | 1.058731119 | 0.93137945 | 0.965874986 |
| Glucose | 1.001611154 | 0.928639184 | 1.080317223 | 0.966728658 | 0.98214648 |
| Lactate | 0.979743002 | 0.929218652 | 1.033014508 | 0.448706571 | 0.575440488 |
| Pyruvate | 1.024806775 | 0.969510545 | 1.083256836 | 0.386571021 | 0.515428028 |
| ***Amino acids*** | | | | | |
| Alanine | 0.911403231 | 0.86390939 | 0.96150807 | 0.000680057 | 0.003264275 |
| Glutamine | 0.955964781 | 0.906796949 | 1.007798565 | 0.094600494 | 0.167293505 |
| Glycine | 0.967486877 | 0.909896372 | 1.028722486 | 0.291150049 | 0.425332246 |
| Histidine | 0.880899271 | 0.836241985 | 0.927941361 | 1.78E-06 | 2.29E-05 |
| Isoleucine | 0.954263072 | 0.901606545 | 1.009994898 | 0.105974731 | 0.185455778 |
| Leucine | 0.935889624 | 0.88351975 | 0.991363678 | 0.024121261 | 0.054031626 |
| Phenylalanine | 1.027123695 | 0.973020616 | 1.084235078 | 0.332375721 | 0.473212891 |
| Total Concentration of Branched-Chain Amino Acids (Leucine + Isoleucine + Valine) | 0.923321054 | 0.870702962 | 0.979118949 | 0.007702478 | 0.023107433 |
| Tyrosine | 1.022975589 | 0.968973341 | 1.079987457 | 0.411689825 | 0.544597564 |
| Valine | 0.908805591 | 0.856746509 | 0.964027975 | 0.001487085 | 0.006245756 |
| ***Apolipoproteins*** | | | | | |
| Apolipoprotein A1 | 0.923044168 | 0.834171239 | 1.021385652 | 0.121068314 | 0.207545681 |
| Apolipoprotein B | 1.088779752 | 0.951457491 | 1.245921504 | 0.216252046 | 0.339535922 |
| **Lipoprotein particle sizes** | | | | | |
| Average Diameter for HDL Particles | 1.272404841 | 1.149021556 | 1.409037167 | 3.67E-06 | 3.63E-05 |
| Average Diameter for LDL Particles | 1.036726136 | 0.981069365 | 1.095540356 | 0.200154707 | 0.317226328 |
| Average Diameter for VLDL Particles | 0.800892882 | 0.741052685 | 0.865565189 | 2.10E-08 | 7.05E-07 |
| ***Other lipids*** | | | | | |
| Phosphatidylcholines | 0.977742448 | 0.896856753 | 1.065923061 | 0.609416845 | 0.705574994 |
| Phosphoglycerides | 0.98176107 | 0.902038647 | 1.068529382 | 0.670112426 | 0.745555547 |
| Sphingomyelins | 1.138388659 | 1.032877327 | 1.254678272 | 0.009006508 | 0.026087818 |
| Total Cholines | 0.994560287 | 0.909772744 | 1.087249724 | 0.90450058 | 0.952907508 |
| ***Fatty acids*** | | | | | |
| Degree of Unsaturation | 0.929148314 | 0.873252375 | 0.988622092 | 0.020262422 | 0.047944886 |
| Docosahexaenoic Acid | 0.900932026 | 0.850088094 | 0.954816945 | 0.000431604 | 0.002132631 |
| Linoleic Acid | 0.949911007 | 0.880711674 | 1.024547474 | 0.183004453 | 0.295622579 |
| Monounsaturated Fatty Acids | 1.010408587 | 0.932592592 | 1.094717588 | 0.800083227 | 0.861851294 |
| Omega-3 Fatty Acids | 0.877071696 | 0.82665882 | 0.930558945 | 1.41E-05 | 0.000107408 |
| Omega-6 Fatty Acids | 0.971247405 | 0.894796286 | 1.054230485 | 0.48552472 | 0.604208541 |
| Polyunsaturated Fatty Acids | 0.920620499 | 0.847615516 | 0.99991339 | 0.049760309 | 0.100719662 |
| Saturated Fatty Acids | 0.99851887 | 0.923192887 | 1.079990917 | 0.97045426 | 0.98214648 |
| Total Fatty Acids | 0.975305862 | 0.897741072 | 1.059572246 | 0.55426656 | 0.669904907 |
| ***Lipoprotein particle concentrations*** | | | | | |
| Concentration of HDL Particles | 0.874860938 | 0.802793585 | 0.953397828 | 0.002303713 | 0.008579408 |
| Concentration of LDL Particles | 1.035522158 | 0.912633875 | 1.174957635 | 0.588117051 | 0.695800454 |
| Concentration of VLDL Particles | 1.034360599 | 0.928318278 | 1.152516195 | 0.540428051 | 0.66271469 |
| Total Concentration of Lipoprotein Particles | 0.889441713 | 0.816021697 | 0.969467557 | 0.007689776 | 0.023107433 |
| ***Lipoprotein particles*** | | | | | |
| Concentration of Chylomicrons and Extremely Large VLDL Particles | 0.966604493 | 0.889539127 | 1.050346431 | 0.422991844 | 0.555176796 |
| Concentration of IDL Particles | 1.182893081 | 1.060483598 | 1.319432043 | 0.00258155 | 0.009035423 |
| Concentration of Large HDL Particles | 1.190495015 | 1.066671322 | 1.328692683 | 0.001859514 | 0.007265079 |
| Concentration of Large LDL Particles | 1.069176953 | 0.952507115 | 1.200137342 | 0.256539813 | 0.384809719 |
| Concentration of Large VLDL Particles | 0.840441532 | 0.765901842 | 0.922235631 | 0.000244076 | 0.001281398 |
| Concentration of Medium HDL Particles | 0.894431108 | 0.813849741 | 0.982991044 | 0.020552722 | 0.047956351 |
| Concentration of Medium LDL Particles | 0.954636432 | 0.860076404 | 1.059592744 | 0.383035732 | 0.515428028 |
| Concentration of Medium VLDL Particles | 0.907798704 | 0.813316551 | 1.013256751 | 0.084509378 | 0.154321474 |
| Concentration of Small HDL Particles | 0.86734677 | 0.815157211 | 0.922877708 | 6.96E-06 | 5.85E-05 |
| Concentration of Small LDL Particles | 1.027397236 | 0.908933131 | 1.16130114 | 0.665445874 | 0.745299379 |
| Concentration of Small VLDL Particles | 0.972269879 | 0.8799775 | 1.074241919 | 0.580515907 | 0.691678528 |
| Concentration of Very Large HDL Particles | 1.22138909 | 1.123752431 | 1.327508861 | 2.54E-06 | 3.05E-05 |
| Concentration of Very Large VLDL Particles | 0.867448071 | 0.788208324 | 0.954653907 | 0.003620586 | 0.012413437 |
| Concentration of Very Small VLDL Particles | 1.313222832 | 1.192995701 | 1.445566155 | 2.66E-08 | 7.46E-07 |
| ***Cholesterol*** | | | | | |
| Clinical LDL Cholesterol | 0.963073747 | 0.86123029 | 1.076960544 | 0.509384179 | 0.62923928 |

| | | | | | |
|---|---|---|---|---|---|
| HDL Cholesterol | 0.983117528 | 0.862805234 | 1.120206549 | 0.798226663 | 0.861851294 |
| LDL Cholesterol | 0.919030537 | 0.824954316 | 1.023835031 | 0.125411313 | 0.212819197 |
| Remnant Cholesterol (Non-HDL, Non-LDL -Cholesterol) | 1.12236158 | 0.988775021 | 1.273996095 | 0.074201075 | 0.136986601 |
| Total Cholesterol | 1.012975715 | 0.905043535 | 1.133779493 | 0.822539718 | 0.88016989 |
| Total Cholesterol Minus HDL-C | 0.997665427 | 0.88183687 | 1.128707971 | 0.97038922 | 0.98214648 |
| VLDL Cholesterol | 1.070823883 | 0.951959156 | 1.204530448 | 0.254346563 | 0.384809719 |
| Cholesterol in Chylomicrons and Extremely Large VLDL | 0.974205737 | 0.895132538 | 1.060264015 | 0.545135031 | 0.663642646 |
| Cholesterol in IDL | 1.092108265 | 0.979690651 | 1.217425584 | 0.111890333 | 0.193789443 |
| Cholesterol in Large HDL | 1.200047234 | 1.07346579 | 1.341554968 | 0.001343718 | 0.005788324 |
| Cholesterol in Large LDL | 0.948249357 | 0.850300276 | 1.057481538 | 0.339455097 | 0.479230726 |
| Cholesterol in Large VLDL | 0.871365166 | 0.784192231 | 0.968228482 | 0.010457577 | 0.029281215 |
| Cholesterol in Medium HDL | 0.873366808 | 0.788669985 | 0.967159389 | 0.009280258 | 0.026425143 |
| Cholesterol in Medium LDL | 0.874441271 | 0.788589929 | 0.969638983 | 0.010936348 | 0.030119778 |
| Cholesterol in Medium VLDL | 1.088493236 | 0.967928513 | 1.224075446 | 0.156856136 | 0.257348237 |
| Cholesterol in Small HDL | 0.867933783 | 0.816125572 | 0.923030815 | 6.47E-06 | 5.72E-05 |
| Cholesterol in Small LDL | 0.894271951 | 0.798839344 | 1.001105328 | 0.052285282 | 0.103340322 |
| Cholesterol in Small VLDL | 1.130382272 | 1.00033932 | 1.277330656 | 0.04936742 | 0.100719662 |
| Cholesterol in Very Large HDL | 1.222660193 | 1.12319544 | 1.330933064 | 3.43E-06 | 3.60E-05 |
| Cholesterol in Very Large VLDL | 0.90447038 | 0.81163853 | 1.007919951 | 0.069188078 | 0.130602215 |
| Cholesterol in Very Small VLDL | 1.341635098 | 1.217212415 | 1.478776189 | 3.25E-09 | 1.93E-07 |
| ***Cholesteryl esters*** | | | | | |
| Cholesteryl Esters in HDL | 0.954151225 | 0.836664537 | 1.08813571 | 0.483889717 | 0.604208541 |
| Cholesteryl Esters in LDL | 0.906219897 | 0.813845533 | 1.009079081 | 0.072622389 | 0.135561794 |
| Cholesteryl Esters in VLDL | 1.155777156 | 1.023106204 | 1.305652169 | 0.019956774 | 0.047896257 |
| Total Esterified Cholesterol | 0.996702747 | 0.892418694 | 1.113172968 | 0.953293278 | 0.98214648 |
| Cholesteryl Esters in Chylomicrons and Extremely Large VLDL | 0.977501231 | 0.896711965 | 1.065569206 | 0.605144184 | 0.705574994 |
| Cholesteryl Esters in IDL | 1.056538128 | 0.951350619 | 1.173355852 | 0.304009072 | 0.440289001 |
| Cholesteryl Esters in Large HDL | 1.199438294 | 1.07076209 | 1.343577844 | 0.001684897 | 0.006803448 |
| Cholesteryl Esters in Large LDL | 0.932044 | 0.837209587 | 1.037620724 | 0.198644826 | 0.317226328 |
| Cholesteryl Esters in Large VLDL | 0.910180256 | 0.816354101 | 1.014790148 | 0.089986992 | 0.161172362 |
| Cholesteryl Esters in Medium HDL | 0.86557398 | 0.781448424 | 0.958755937 | 0.005651223 | 0.01791331 |
| Cholesteryl Esters in Medium LDL | 0.871357571 | 0.786046291 | 0.965927866 | 0.008808907 | 0.025963095 |
| Cholesteryl Esters in Medium VLDL | 1.147171381 | 1.025264827 | 1.28357293 | 0.016609313 | 0.04200548 |
| Cholesteryl Esters in Small HDL | 0.856815924 | 0.804636019 | 0.912379648 | 1.43E-06 | 2.01E-05 |
| Cholesteryl Esters in Small LDL | 0.893874769 | 0.796654895 | 1.002958883 | 0.056175365 | 0.108476567 |
| Cholesteryl Esters in Small VLDL | 1.164214216 | 1.031645076 | 1.313818844 | 0.013698979 | 0.035959821 |
| Cholesteryl Esters in Very Large HDL | 1.238036148 | 1.127107428 | 1.359882355 | 8.26E-06 | 6.61E-05 |
| Cholesteryl Esters in Very Large VLDL | 0.918100556 | 0.817055288 | 1.031642097 | 0.150909935 | 0.251018506 |
| Cholesteryl Esters in Very Small VLDL | 1.319485087 | 1.197765422 | 1.453574183 | 1.97E-08 | 7.05E-07 |
| ***Free cholesterol*** | | | | | |
| Free Cholesterol in HDL | 1.066475389 | 0.950434791 | 1.19668363 | 0.27350425 | 0.403058895 |
| Free Cholesterol in LDL | 0.962806024 | 0.867080964 | 1.069099055 | 0.478070084 | 0.603878001 |
| Free Cholesterol in VLDL | 0.981688564 | 0.880825909 | 1.09410092 | 0.738295312 | 0.816010608 |
| Total Free Cholesterol | 1.057229238 | 0.941057432 | 1.18774224 | 0.348733136 | 0.484191461 |
| Free Cholesterol in Chylomicrons and Extremely Large VLDL | 0.968118867 | 0.892328654 | 1.050346344 | 0.435985067 | 0.567794506 |
| Free Cholesterol in IDL | 1.18641767 | 1.06246228 | 1.324834693 | 0.00239648 | 0.008579408 |
| Free Cholesterol in Large HDL | 1.189540566 | 1.070679428 | 1.321597035 | 0.001231697 | 0.005445396 |
| Free Cholesterol in Large LDL | 1.000120857 | 0.897477241 | 1.114503726 | 0.998254773 | 0.998254773 |
| Free Cholesterol in Large VLDL | 0.844756119 | 0.763360708 | 0.934830537 | 0.001100021 | 0.00499469 |
| Free Cholesterol in Medium HDL | 0.911592302 | 0.826581732 | 1.005345863 | 0.063851113 | 0.121897579 |
| Free Cholesterol in Medium LDL | 0.907556814 | 0.8228822 | 1.000944448 | 0.052249386 | 0.103340322 |
| Free Cholesterol in Medium VLDL | 0.985110801 | 0.877018891 | 1.106524957 | 0.800290487 | 0.861851294 |
| Free Cholesterol in Small HDL | 0.900401839 | 0.838349725 | 0.967046864 | 0.003980429 | 0.013374241 |
| Free Cholesterol in Small LDL | 0.934594114 | 0.850868304 | 1.026558579 | 0.157778979 | 0.257348237 |
| Free Cholesterol in Small VLDL | 1.059170263 | 0.939249236 | 1.194402512 | 0.348416135 | 0.484191461 |
| Free Cholesterol in Very Large HDL | 1.196661337 | 1.113874136 | 1.285601586 | 9.19E-07 | 1.40E-05 |
| Free Cholesterol in Very Large VLDL | 0.899724632 | 0.814250518 | 0.994171198 | 0.038010036 | 0.081025008 |
| Free Cholesterol in Very Small VLDL | 1.347583905 | 1.225262884 | 1.482116536 | 8.03E-10 | 1.35E-07 |
| ***Phospholipids*** | | | | | |
| Phospholipids in HDL | 0.960422818 | 0.866712985 | 1.064264647 | 0.440757144 | 0.569593848 |

| | | | | | |
|---|---|---|---|---|---|
| Phospholipids in LDL | 0.935142782 | 0.834734631 | 1.04762878 | 0.247239309 | 0.377601854 |
| Phospholipids in VLDL | 0.970097411 | 0.874127799 | 1.076603431 | 0.567862233 | 0.68143468 |
| Total Phospholipids in Lipoprotein Particles | 1.000718555 | 0.910546492 | 1.099820422 | 0.988104774 | 0.994021569 |
| Phospholipids in Chylomicrons and Extremely Large VLDL | 0.980626453 | 0.900263667 | 1.06816289 | 0.65382962 | 0.737203866 |
| Phospholipids in IDL | 1.31485828 | 1.169724009 | 1.478000181 | 4.50E-06 | 4.20E-05 |
| Phospholipids in Large HDL | 1.125179934 | 1.006493346 | 1.257862149 | 0.038101045 | 0.081025008 |
| Phospholipids in Large LDL | 0.983328503 | 0.878682302 | 1.100437488 | 0.769639956 | 0.845094854 |
| Phospholipids in Large VLDL | 0.833292222 | 0.740817767 | 0.937310036 | 0.002376084 | 0.008579408 |
| Phospholipids in Medium HDL | 0.901952489 | 0.828826226 | 0.981530588 | 0.016752185 | 0.04200548 |
| Phospholipids in Medium LDL | 0.864692863 | 0.777722789 | 0.961388503 | 0.007187737 | 0.022361847 |
| Phospholipids in Medium VLDL | 0.937304447 | 0.837018577 | 1.049605887 | 0.262108933 | 0.389684078 |
| Phospholipids in Small HDL | 0.887032525 | 0.83276715 | 0.944833981 | 0.000197811 | 0.001107739 |
| Phospholipids in Small LDL | 0.948767235 | 0.853297706 | 1.054918184 | 0.331085519 | 0.473212891 |
| Phospholipids in Small VLDL | 1.006348677 | 0.897034203 | 1.128984444 | 0.914099599 | 0.95384306 |
| Phospholipids in Very Large HDL | 1.221151606 | 1.114937363 | 1.337484324 | 1.68E-05 | 0.000117755 |
| Phospholipids in Very Large VLDL | 0.877997465 | 0.785037391 | 0.981965391 | 0.022685273 | 0.05214766 |
| Phospholipids in Very Small VLDL | 1.307179485 | 1.196035786 | 1.428651404 | 3.45E-09 | 1.93E-07 |
| ***Total lipids*** | | | | | |
| Total Lipids in HDL | 0.971495066 | 0.868457059 | 1.086758008 | 0.613178268 | 0.705574994 |
| Total Lipids in LDL | 0.935020385 | 0.836760962 | 1.044818244 | 0.23561323 | 0.366509469 |
| Total Lipids in Lipoprotein Particles | 0.956013999 | 0.867574666 | 1.053468712 | 0.363747235 | 0.498790333 |
| Total Lipids in VLDL | 0.905452419 | 0.823348259 | 0.995743993 | 0.040569156 | 0.085195228 |
| Total Lipids in Chylomicrons and Extremely Large VLDL | 0.942295677 | 0.871450998 | 1.01889968 | 0.136106049 | 0.228658162 |
| Total Lipids in IDL | 1.17986274 | 1.051316766 | 1.324126212 | 0.004950452 | 0.016307373 |
| Total Lipids in Large HDL | 1.170940663 | 1.047508251 | 1.308917648 | 0.005492681 | 0.017745584 |
| Total Lipids in Large LDL | 0.973027319 | 0.871104727 | 1.086875246 | 0.628148876 | 0.713033859 |
| Total Lipids in Large VLDL | 0.819589353 | 0.748933971 | 0.896910455 | 1.52E-05 | 0.000111255 |
| Total Lipids in Medium HDL | 0.89779155 | 0.821521813 | 0.981142138 | 0.017300408 | 0.042742184 |
| Total Lipids in Medium LDL | 0.877844695 | 0.789779375 | 0.975729847 | 0.015714473 | 0.04061587 |
| Total Lipids in Medium VLDL | 0.855728273 | 0.773835404 | 0.946287639 | 0.002400192 | 0.008579408 |
| Total Lipids in Small HDL | 0.879577208 | 0.826187375 | 0.936417195 | 5.92E-05 | 0.000382233 |
| Total Lipids in Small LDL | 0.904839717 | 0.806005965 | 1.015792623 | 0.090179774 | 0.161172362 |
| Total Lipids in Small VLDL | 0.963192803 | 0.870191613 | 1.066133439 | 0.469145242 | 0.597093945 |
| Total Lipids in Very Large HDL | 1.214528886 | 1.118946554 | 1.318276025 | 3.36E-06 | 3.60E-05 |
| Total Lipids in Very Large VLDL | 0.855911747 | 0.780804175 | 0.938244111 | 0.000899175 | 0.004196148 |
| Total Lipids in Very Small VLDL | 1.290119726 | 1.1771051 | 1.41398496 | 5.15E-08 | 9.83E-07 |
| ***Triglycerides*** | | | | | |
| Total Triglycerides | 0.900817687 | 0.829473402 | 0.978298403 | 0.013096528 | 0.034924074 |
| Triglycerides in HDL | 1.017929424 | 0.948542684 | 1.092391866 | 0.621768727 | 0.710592831 |
| Triglycerides in LDL | 1.097374788 | 1.012912642 | 1.188879846 | 0.022969803 | 0.05214766 |
| Triglycerides in VLDL | 0.854862745 | 0.787377267 | 0.92813235 | 0.000185831 | 0.00107654 |
| Triglycerides in Chylomicrons and Extremely Large VLDL | 0.922659346 | 0.850379801 | 1.001082419 | 0.053116581 | 0.103762623 |
| Triglycerides in IDL | 1.151118426 | 1.067254922 | 1.241571815 | 0.000265875 | 0.001353545 |
| Triglycerides in Large HDL | 1.070485185 | 1.00028988 | 1.145606443 | 0.049028944 | 0.100719662 |
| Triglycerides in Large LDL | 1.13302194 | 1.048001599 | 1.224939655 | 0.001700862 | 0.006803448 |
| Triglycerides in Large VLDL | 0.798536415 | 0.737245644 | 0.864922583 | 3.36E-08 | 8.07E-07 |
| Triglycerides in Medium HDL | 1.00424679 | 0.936840823 | 1.076502635 | 0.9048438 | 0.952907508 |
| Triglycerides in Medium LDL | 1.049859881 | 0.968243227 | 1.138356293 | 0.238641815 | 0.367814908 |
| Triglycerides in Medium VLDL | 0.812305485 | 0.74778668 | 0.882390952 | 8.52E-07 | 1.40E-05 |
| Triglycerides in Small HDL | 0.961954923 | 0.884492325 | 1.046201587 | 0.36518578 | 0.498790333 |
| Triglycerides in Small LDL | 0.978227434 | 0.900727671 | 1.062395375 | 0.601169557 | 0.705574994 |
| Triglycerides in Small VLDL | 0.903182873 | 0.834150576 | 0.977928116 | 0.012068525 | 0.03270181 |
| Triglycerides in Very Large HDL | 1.08026321 | 1.007063854 | 1.158783127 | 0.031037816 | 0.068609908 |
| Triglycerides in Very Large VLDL | 0.837132508 | 0.769565657 | 0.910631639 | 3.47E-05 | 0.000233121 |
| Triglycerides in Very Small VLDL | 1.08741982 | 1.006260841 | 1.175124597 | 0.034202942 | 0.074624602 |
| ***Ketone bodies*** | | | | | |
| 3-Hydroxybutyrate | 1.121536533 | 1.05968157 | 1.187002049 | 7.41E-05 | 0.000461161 |
| Acetate | 1.00328899 | 0.949211615 | 1.060447198 | 0.90753096 | 0.952907508 |
| Acetoacetate | 1.112966708 | 1.051589391 | 1.177926388 | 0.000217321 | 0.001177739 |
| Acetone | 1.10617439 | 1.051109787 | 1.164123669 | 0.000107367 | 0.000644201 |
| ***Fluid balance*** | | | | | |

| | | | | | |
|---|---|---|---|---|---|
| Albumin | 0.861279777 | 0.816181232 | 0.908870266 | 5.27E-08 | 9.83E-07 |
| Creatinine | 0.97238077 | 0.913009788 | 1.035612513 | 0.383576061 | 0.515428028 |
| ***Inflammation*** | | | | | |
| Glycoprotein Acetyls | 1.078753507 | 1.012418344 | 1.149435048 | 0.019226147 | 0.046811488 |

**Supplementary Table 4: Results of per-metabolite COX-regressions after adjustment for PCP-HF characteristics.** P-values were adjusted following the Benjamini-Hochberg method. HR = Hazard ratio. CI = Confidence interval.

| Characteristic | Validation | Derivation | Whole Cohort, n=68,327 |
|---|---|---|---|
| ***Sociodemographics*** | | | |
| Age at recruitment (years) | 57 (13) | 57 (13) | 57 (13) |
| Sex | | | |
| Female | 7,565 (55.373 %) | 30,582 (55.961 %) | 38,147 (55.843 %) |
| Male | 6,097 (44.627 %) | 24,067 (44.039 %) | 30,164 (44.157 %) |
| Ethnicity | | | |
| Black | 184 (1.347 %) | 699 (1.279 %) | 883 (1.293 %) |
| White | 13,478 (98.653 %) | 53,950 (98.721 %) | 67,428 (98.707 %) |
| Higher Education | 4,574 (33.480 %) | 18,408 (33.684 %) | 22,982 (33.643 %) |
| ***Lifestyle factors*** | | | |
| Current smoker | 1,401 (10.255 %) | 5,586 (10.222 %) | 6,987 (10.228 %) |
| Alcohol consumption | 12,778 (93.529 %) | 51,166 (93.627 %) | 63,944 (93.607 %) |
| Daily physical activity (minutes) | 100 (120) | 100 (120.75) | 100 (120) |
| ***Clinical chemistry*** | | | |
| Total cholesterol (mmol $l^{-1}$) | 5.885 (1.385) | 5.887 (1.39) | 5.887 (1.389) |
| HDL cholesterol (mmol $l^{-1}$) | 1.415 (0.486) | 1.422 (0.487) | 1.421 (0.486) |
| LDL cholesterol (mmol $l^{-1}$) | 3.723 (1.054) | 3.716 (1.069) | 3.718 (1.066) |
| Triglycerides (mmol $l^{-1}$) | 1.489 (1.075) | 1.479 (1.037) | 1.481 (1.044) |
| Glucose (mmol $l^{-1}$) | 4.898 (0.672) | 4.9 (0.667) | 4.899 (0.667) |
| Glycated hemoglobin (%) | 3.47 (0.46) | 3.47 (0.47) | 3.47 (0.47) |
| Creatinine (µmol $l^{-1}$) | 70 (18.6) | 70.2 (18.9) | 70.1 (18.9) |
| C-reactive protein (mg $l^{-1}$) | 1.34 (2.07) | 1.32 (2.06) | 1.32 (2.06) |
| ***Physical measurements*** | | | |
| SBP (mmHg) | 132 (24) | 132 (24) | 132 (24) |
| BMI ($m^2$/kg) | 26.504 (5.527) | 26.471 (5.501) | 26.480 (5.513) |
| Waist/hip ratio | 0.865 (0.127) | 0.864 (0.128) | 0.864 (0.128) |
| ***Past medical history*** | | | |
| Treatment for Diabetes | 72 (0.527 %) | 303 (0.554 %) | 375 (0.549 %) |
| Treatment for Hypertension | 1,780 (13.029 %) | 7,297 (13.352 %) | 9,077 (13.288 %) |
| Family history of heart disease | 5,631 (41.217 %) | 21,993 (40.244 %) | 27,624 (40.44 %) |
| ***Incident Endpoints*** | | | |
| Heart Failure | 292 (2.137 %) | 1,168 (2.137 %) | 1,460 (2.137 %) |
| Hypertension | 4,179 (30.588 %) | 16,666 (30.496 %) | 20,845 (30.515 %) |
| Coronary artery disease | 464 (3.396 %) | 1,873 (3.427 %) | 2,337 (3.421 %) |

**Supplementary table 5: Population characteristics of validation and partition splits.** Listed clinical chemistry measurements were not 1H-NMR-derived. Median (IQR) for continuous, *n* (%) for categorical variables. Mann-Whitney-U for continuous, Chi-Square for categorical variables; BH-correction for multiple testing. No characteristic reached the significance threshold of $P < 0.01$.

| Features | Age & Sex | Age & Sex<br>+ Metabolomics | PCP-HF | PCP-HF<br>+ Metabolomics |
|---|---|---|---|---|
| Age | 0.57038494 | 0.684807263 | 0.583318536 | 0.654127312 |
| Sex | 0.43309518 | 0.478545595 | 0.395481221 | 0.49104184 |
| Ethnicity | 0 | 0 | -0.225385012 | -0.141438353 |
| Smoking status | 0 | 0 | 0.603802991 | 0.558646426 |
| Cholesterol | 0 | 0 | -0.01927186 | 0 |
| HDL | 0 | 0 | -0.069066718 | 0 |
| LDL | 0 | 0 | 0 | 0 |
| Glucose (non-NMR) | 0 | 0 | 0.076751634 | 0.052158513 |
| SBP | 0 | 0 | 0.084016998 | 0.056846869 |
| BMI | 0 | 0 | 0.249623209 | 0.253182603 |
| DM Medication | 0 | 0 | 0.406307206 | 0.294952307 |
| HT Medication | 0 | 0 | 0.570218175 | 0.555701805 |
| ***Metabolites*** | | | | |
| ***Glycolysis related metabolites*** | | | | |
| Citrate | 0 | 0 | 0 | 0 |
| Glucose | 0 | 0.07163374 | 0 | 0.00933464 |
| Lactate | 0 | 0 | 0 | 0 |
| Pyruvate | 0 | 0 | 0 | 0 |
| ***Amino acids*** | | | | |
| Alanine | 0 | -0.016664873 | 0 | -0.004138383 |
| Glutamine | 0 | 0 | 0 | 0 |
| Glycine | 0 | -0.053644761 | 0 | -0.002154942 |
| Histidine | 0 | -0.055812549 | 0 | -0.031552997 |
| Isoleucine | 0 | 0 | 0 | 0 |
| Leucine | 0 | 0 | 0 | 0 |
| Phenylalanine | 0 | 0.043283948 | 0 | 0.027938961 |
| Total Concentration of Branched-Chain Amino Acids (Leucine + Isoleucine + Valine) | 0 | 0 | 0 | 0 |
| Tyrosine | 0 | 0.045237438 | 0 | 0.002394527 |
| Valine | 0 | 0 | 0 | 0 |
| ***Apolipoproteins*** | | | | |
| Apolipoprotein A1 | 0 | 0 | 0 | 0 |
| Apolipoprotein B | 0 | 0 | 0 | 0 |
| ***Lipoprotein particle sizes*** | | | | |
| Average Diameter for HDL Particles | 0 | 0 | 0 | 0 |
| Average Diameter for LDL Particles | 0 | 0 | 0 | 0 |
| Average Diameter for VLDL Particles | 0 | 0 | 0 | 0 |
| ***Other lipids*** | | | | |
| Phosphatidylcholines | 0 | 0 | 0 | 0 |
| Phosphoglycerides | 0 | 0 | 0 | 0 |
| Sphingomyelins | 0 | 0 | 0 | 0 |
| Total Cholines | 0 | 0 | 0 | 0 |
| ***Fatty acids*** | | | | |
| Degree of Unsaturation | 0 | 0 | 0 | 0 |
| Docosahexaenoic Acid | 0 | -0.0296148 | 0 | 0 |
| Linoleic Acid | 0 | -0.1196912 | 0 | 0 |
| Monounsaturated Fatty Acids | 0 | 0 | 0 | 0 |
| Omega-3 Fatty Acids | 0 | -0.1503239 | 0 | -0.0983334 |
| Omega-6 Fatty Acids | 0 | 0 | 0 | 0 |
| Polyunsaturated Fatty Acids | 0 | 0 | 0 | 0 |
| Saturated Fatty Acids | 0 | 0 | 0 | 0 |
| Total Fatty Acids | 0 | 0 | 0 | 0 |
| ***Lipoprotein particle concentrations*** | | | | |
| Concentration of HDL Particles | 0 | 0 | 0 | 0 |
| Concentration of LDL Particles | 0 | 0 | 0 | 0 |
| Concentration of VLDL Particles | 0 | 0 | 0 | 0 |
| Total Concentration of Lipoprotein Particles | 0 | 0 | 0 | 0 |
| Concentration of Chylomicrons and Extremely Large VLDL Particles | 0 | 0 | 0 | 0 |
| Concentration of IDL Particles | 0 | 0 | 0 | 0 |
| Concentration of Large HDL Particles | 0 | 0 | 0 | 0 |

| | | | | |
|---|---|---|---|---|
| Concentration of Large LDL Particles | 0 | 0 | 0 | 0 |
| Concentration of Large VLDL Particles | 0 | 0 | 0 | 0 |
| Concentration of Medium HDL Particles | 0 | 0 | 0 | 0 |
| Concentration of Medium LDL Particles | 0 | 0 | 0 | 0 |
| Concentration of Medium VLDL Particles | 0 | 0 | 0 | 0 |
| Concentration of Small HDL Particles | 0 | 0 | 0 | 0 |
| Concentration of Small LDL Particles | 0 | 0 | 0 | 0 |
| Concentration of Small VLDL Particles | 0 | 0 | 0 | 0 |
| Concentration of Very Large HDL Particles | 0 | 0 | 0 | 0 |
| Concentration of Very Large VLDL Particles | 0 | 0 | 0 | 0 |
| Concentration of Very Small VLDL Particles | 0 | 0 | 0 | 0 |
| ***Cholesterol*** | | | | |
| Clinical LDL Cholesterol | 0 | 0 | 0 | 0 |
| HDL Cholesterol | 0 | 0 | 0 | 0 |
| LDL Cholesterol | 0 | 0 | 0 | 0 |
| Remnant Cholesterol (Non-HDL, Non-LDL -Cholesterol) | 0 | 0 | 0 | 0 |
| Total Cholesterol | 0 | 0 | 0 | 0 |
| Total Cholesterol Minus HDL-C | 0 | 0 | 0 | 0 |
| VLDL Cholesterol | 0 | 0 | 0 | 0 |
| Cholesterol in Chylomicrons and Extremely Large VLDL | 0 | 0 | 0 | 0 |
| Cholesterol in IDL | 0 | 0 | 0 | 0 |
| Cholesterol in Large HDL | 0 | 0 | 0 | 0 |
| Cholesterol in Large LDL | 0 | 0 | 0 | 0 |
| Cholesterol in Large VLDL | 0 | 0 | 0 | 0 |
| Cholesterol in Medium HDL | 0 | -0.002127536 | 0 | 0 |
| Cholesterol in Medium LDL | 0 | 0 | 0 | 0 |
| Cholesterol in Medium VLDL | 0 | 0 | 0 | 0 |
| Cholesterol in Small HDL | 0 | 0 | 0 | 0 |
| Cholesterol in Small LDL | 0 | 0 | 0 | 0 |
| Cholesterol in Small VLDL | 0 | 0 | 0 | 0 |
| Cholesterol in Very Large HDL | 0 | 0 | 0 | 0 |
| Cholesterol in Very Large VLDL | 0 | 0 | 0 | 0 |
| Cholesterol in Very Small VLDL | 0 | 0 | 0 | 0 |
| ***Cholesteryl esters*** | | | | |
| Cholesteryl Esters in HDL | 0 | 0 | 0 | 0 |
| Cholesteryl Esters in LDL | 0 | 0 | 0 | 0 |
| Cholesteryl Esters in VLDL | 0 | 0 | 0 | 0 |
| Total Esterified Cholesterol | 0 | 0 | 0 | 0 |
| Cholesteryl Esters in Chylomicrons and Extremely Large VLDL | 0 | 0 | 0 | 0 |
| Cholesteryl Esters in IDL | 0 | 0 | 0 | 0 |
| Cholesteryl Esters in Large HDL | 0 | 0 | 0 | 0 |
| Cholesteryl Esters in Large LDL | 0 | 0 | 0 | 0 |
| Cholesteryl Esters in Large VLDL | 0 | 0 | 0 | 0 |
| Cholesteryl Esters in Medium HDL | 0 | -0.000684677 | 0 | 0 |
| Cholesteryl Esters in Medium LDL | 0 | 0 | 0 | 0 |
| Cholesteryl Esters in Medium VLDL | 0 | 0 | 0 | 0 |
| Cholesteryl Esters in Small HDL | 0 | -0.035098357 | 0 | -0.091599835 |
| Cholesteryl Esters in Small LDL | 0 | 0 | 0 | 0 |
| Cholesteryl Esters in Small VLDL | 0 | 0 | 0 | 0 |
| Cholesteryl Esters in Very Large HDL | 0 | 0 | 0 | 0 |
| Cholesteryl Esters in Very Large VLDL | 0 | 0 | 0 | 0 |
| Cholesteryl Esters in Very Small VLDL | 0 | 0 | 0 | 0 |
| ***Free cholesterol*** | | | | |
| Free Cholesterol in HDL | 0 | 0 | 0 | 0 |
| Free Cholesterol in LDL | 0 | 0 | 0 | 0 |
| Free Cholesterol in VLDL | 0 | 0 | 0 | 0 |
| Total Free Cholesterol | 0 | 0 | 0 | 0 |
| Free Cholesterol in Chylomicrons and Extremely Large VLDL | 0 | 0 | 0 | 0 |
| Free Cholesterol in IDL | 0 | 0 | 0 | 0 |
| Free Cholesterol in Large HDL | 0 | 0 | 0 | 0 |

| | | | | |
|---|---|---|---|---|
| Free Cholesterol in Large LDL | 0 | 0 | 0 | 0 |
| Free Cholesterol in Large VLDL | 0 | 0 | 0 | 0 |
| Free Cholesterol in Medium HDL | 0 | -0.006816866 | 0 | 0 |
| Free Cholesterol in Medium LDL | 0 | 0 | 0 | 0 |
| Free Cholesterol in Medium VLDL | 0 | 0 | 0 | 0 |
| Free Cholesterol in Small HDL | 0 | 0 | 0 | 0 |
| Free Cholesterol in Small LDL | 0 | 0 | 0 | 0 |
| Free Cholesterol in Small VLDL | 0 | 0 | 0 | 0 |
| Free Cholesterol in Very Large HDL | 0 | 0 | 0 | 0 |
| Free Cholesterol in Very Large VLDL | 0 | 0 | 0 | 0 |
| Free Cholesterol in Very Small VLDL | 0 | 0 | 0 | 0 |
| ***Phospholipids*** | | | | |
| Phospholipids in HDL | 0 | 0 | 0 | 0 |
| Phospholipids in LDL | 0 | 0 | 0 | 0 |
| Phospholipids in VLDL | 0 | 0 | 0 | 0 |
| Total Phospholipids in Lipoprotein Particles | 0 | 0 | 0 | 0 |
| Phospholipids in Chylomicrons and Extremely Large VLDL | 0 | 0 | 0 | 0 |
| Phospholipids in IDL | 0 | 0 | 0 | 0 |
| Phospholipids in Large HDL | 0 | 0 | 0 | 0 |
| Phospholipids in Large LDL | 0 | 0 | 0 | 0 |
| Phospholipids in Large VLDL | 0 | 0 | 0 | 0 |
| Phospholipids in Medium HDL | 0 | 0 | 0 | 0 |
| Phospholipids in Medium LDL | 0 | 0 | 0 | 0 |
| Phospholipids in Medium VLDL | 0 | 0 | 0 | 0 |
| Phospholipids in Small HDL | 0 | 0 | 0 | 0 |
| Phospholipids in Small LDL | 0 | 0 | 0 | 0 |
| Phospholipids in Small VLDL | 0 | 0 | 0 | 0 |
| Phospholipids in Very Large HDL | 0 | 0 | 0 | 0 |
| Phospholipids in Very Large VLDL | 0 | 0 | 0 | 0 |
| Phospholipids in Very Small VLDL | 0 | 0.100427729 | 0 | 0.064102359 |
| ***Total lipids*** | | | | |
| Total Lipids in HDL | 0 | 0 | 0 | 0 |
| Total Lipids in LDL | 0 | 0 | 0 | 0 |
| Total Lipids in Lipoprotein Particles | 0 | 0 | 0 | 0 |
| Total Lipids in VLDL | 0 | 0 | 0 | 0 |
| Total Lipids in Chylomicrons and Extremely Large VLDL | 0 | 0 | 0 | 0 |
| Total Lipids in IDL | 0 | 0 | 0 | 0 |
| Total Lipids in Large HDL | 0 | 0 | 0 | 0 |
| Total Lipids in Large LDL | 0 | 0 | 0 | 0 |
| Total Lipids in Large VLDL | 0 | 0 | 0 | 0 |
| Total Lipids in Medium HDL | 0 | 0 | 0 | 0 |
| Total Lipids in Medium LDL | 0 | 0 | 0 | 0 |
| Total Lipids in Medium VLDL | 0 | 0 | 0 | 0 |
| Total Lipids in Small HDL | 0 | 0 | 0 | 0 |
| Total Lipids in Small LDL | 0 | 0 | 0 | 0 |
| Total Lipids in Small VLDL | 0 | 0 | 0 | 0 |
| Total Lipids in Very Large HDL | 0 | 0 | 0 | 0 |
| Total Lipids in Very Large VLDL | 0 | 0 | 0 | 0 |
| Total Lipids in Very Small VLDL | 0 | 0 | 0 | 0 |
| ***Triglycerides*** | | | | |
| Total Triglycerides | 0 | 0 | 0 | 0 |
| Triglycerides in HDL | 0 | 0 | 0 | 0 |
| Triglycerides in LDL | 0 | 0 | 0 | 0 |
| Triglycerides in VLDL | 0 | 0 | 0 | 0 |
| Triglycerides in Chylomicrons and Extremely Large VLDL | 0 | 0 | 0 | 0 |
| Triglycerides in IDL | 0 | 0.049168029 | 0 | 0.019934548 |
| Triglycerides in Large HDL | 0 | 0 | 0 | 0 |
| Triglycerides in Large LDL | 0 | 0.02708056 | 0 | 0 |
| Triglycerides in Large VLDL | 0 | -0.016556512 | 0 | -0.040824907 |
| Triglycerides in Medium HDL | 0 | 0 | 0 | 0 |
| Triglycerides in Medium LDL | 0 | 0 | 0 | 0 |

| | | | | |
|---|---|---|---|---|
| Triglycerides in Medium VLDL | 0 | 0 | 0 | 0 |
| Triglycerides in Small HDL | 0 | 0 | 0 | 0 |
| Triglycerides in Small LDL | 0 | 0 | 0 | 0 |
| Triglycerides in Small VLDL | 0 | 0 | 0 | 0 |
| Triglycerides in Very Large HDL | 0 | 0 | 0 | 0 |
| Triglycerides in Very Large VLDL | 0 | 0 | 0 | 0 |
| Triglycerides in Very Small VLDL | 0 | 0 | 0 | 0 |
| ***Ketone bodies*** | | | | |
| 3-Hydroxybutyrate | 0 | 0.0857555 | 0 | 0.06340529 |
| Acetate | 0 | 0 | 0 | 0 |
| Acetoacetate | 0 | 0.00554856 | 0 | 0.00603817 |
| Acetone | 0 | 0 | 0 | 0.00911172 |
| ***Fluid balance*** | | | | |
| Albumin | 0 | -0.1383381 | 0 | -0.0962005 |
| Creatinine | 0 | 0 | 0 | 0 |
| ***Inflammation*** | | | | |
| Glycoprotein Acetyls | 0 | 0.16858797 | 0 | 0.0540613 |

**Supplementary Table 6: Final risk model individual-feature coefficients.**

| Models | Alpha | Lambda |
|---|---|---|
| Age & Sex | 0.2 | 0.00526464 |
| Age & Sex + Metabolomics | 0.9 | 0.0008850015 |
| PCP-HF | 0.1 | 0.004152965 |
| PCP-HF + Metabolomics | 0.7 | 0.001248799 |

**Supplementary Table 7: EN-model hyperparameters.** Models were optimized for Harrel's C.

| | Age & Sex | Age & Sex + Metabolomics | PCP-HF | PCP-HF + Metabolomics |
|---|---|---|---|---|
| Harrell's C | 0.713 [0.711-0.714] | 0.746 [0.744-0.747] | 0.756 [0.754-0.757] | 0.766 [0.764-0.767] |
| Sensitivity | 0.700 [0.689-0.711] | 0.743 [0.738-0.748] | 0.739 [0.713-0.765] | 0.748 [0.746-0.750] |
| Specificity | 0.625 [0.613-0.637] | 0.628 [0.625-0.631] | 0.636 [0.610-0.663] | 0.648 [0.646-0.651] |

**Supplementary Table 8: Absolute discriminative capacities in the derivation partition.** Model performance is assessed in the derivation partition, mean and 95% confidence intervals (in square brackets) are calculated from performance within the 10 cross-validation splits. Sensitivity and Specificity are calculated at the Youden point.

| | **Age & Sex** vs. **Age & Sex + Metabolomics** | **Age & Sex** vs. **PCP-HF** | **Age & Sex** vs. **PCP-HF + Metabolomics** | **Age & Sex + Metabolomics** vs. **PCP-HF** | **Age & Sex + Metabolomics** vs. **PCP-HF + Metabolomics** | **PCP-HF** vs. **PCP-HF + Metabolomics** |
|---|---|---|---|---|---|---|
| ***NRI*** | | | | | | |
| Cases | 0.549 [0.546-0.553] | 0.118 [0.112-0.123] | 0.446 [0.440-0.452] | -0.274 [-0.279-(-0.270)] | 0.080 [0.074-0.087] | 0.736 [0.733-0.739] |
| Non-Cases | 0.026 [0.025-0.027] | 0.385 [0.384-0.386] | 0.166 [0.165-0.167] | 0.335 [0.335-0.336] | 0.233 [0.233-0.234] | -0.419 [-0.420-(-0.419)] |
| Overall | 0.575 [0.572-0.579] | 0.503 [0.497-0.508] | 0.612 [0.606-0.618] | 0.061 [0.056-0.065] | 0.314 [0.308-0.320] | 0.317 [0.314-0.320] |
| ***IDI*** | | | | | | |
| Absolute (%) | 0.531 [0.523-0.538] | 0.674 [0.657-0.691] | 0.975 [0.955-0.996] | 0.143 [0.129-0.157] | 0.445 [0.428-0.461] | 0.301 [0.296-0.307] |
| Relative (%) | 90.605 [89.461-91.749] | 115.043 [112.626-117.459] | 166.475 [163.621-169.330] | 12.825 [11.579-14.072] | 39.808[38.455-41.160] | 23.921 [23.519-24.322] |

**Supplementary Table 9: Metrics of relative performance in the derivation partition.** Model performance is assessed in the derivation partition, mean and 95% confidence intervals (in square brackets) are calculated from performance within the 10 cross-validation splits.

| Section/Topic | | | Checklist Item | Page |
|---|---|---|---|---|
| **Title and abstract** | | | | |
| Title | 1 | D;V | Identify the study as developing and/or validating a multivariable prediction model, the target population, and the outcome to be predicted. | 1 |
| Abstract | 2 | D;V | Provide a summary of objectives, study design, setting, participants, sample size, predictors, outcome, statistical analysis, results, and conclusions. | 1 |
| **Introduction** | | | | |
| Background and objectives | 3a | D;V | Explain the medical context (including whether diagnostic or prognostic) and rationale for developing or validating the multivariable prediction model, including references to existing models. | 2 |
| | 3b | D;V | Specify the objectives, including whether the study describes the development or validation of the model or both. | 2 |
| **Methods** | | | | |
| Source of data | 4a | D;V | Describe the study design or source of data (e.g., randomized trial, cohort, or registry data), separately for the development and validation data sets, if applicable. | 3 |
| | 4b | D;V | Specify the key study dates, including start of accrual; end of accrual; and, if applicable, end of follow-up. | 3 |
| Participants | 5a | D;V | Specify key elements of the study setting (e.g., primary care, secondary care, general population) including number and location of centres. | 3 |
| | 5b | D;V | Describe eligibility criteria for participants. | 3 |
| | 5c | D;V | Give details of treatments received, if relevant. | NA |
| Outcome | 6a | D;V | Clearly define the outcome that is predicted by the prediction model, including how and when assessed. | 4 |
| | 6b | D;V | Report any actions to blind assessment of the outcome to be predicted. | N/A |
| Predictors | 7a | D;V | Clearly define all predictors used in developing or validating the multivariable prediction model, including how and when they were measured. | 3, 4 |
| | 7b | D;V | Report any actions to blind assessment of predictors for the outcome and other predictors. | NA |
| Sample size | 8 | D;V | Explain how the study size was arrived at. | 3 |
| Missing data | 9 | D;V | Describe how missing data were handled (e.g., complete-case analysis, single imputation, multiple imputation) with details of any imputation method. | 3, 4 |
| Statistical analysis methods | 10a | D | Describe how predictors were handled in the analyses. | 3, 4, 5 |
| | 10b | D | Specify type of model, all model-building procedures (including any predictor selection), and method for internal validation. | 3, 4, 5 |
| | 10c | V | For validation, describe how the predictions were calculated. | 5 |
| | 10d | D;V | Specify all measures used to assess model performance and, if relevant, to compare multiple models. | 5 |
| | 10e | V | Describe any model updating (e.g., recalibration) arising from the validation, if done. | NA |
| Risk groups | 11 | D;V | Provide details on how risk groups were created, if done. | 5 |
| Development vs. validation | 12 | V | For validation, identify any differences from the development data in setting, eligibility criteria, outcome, and predictors. | NA |
| **Results** | | | | |
| Participants | 13a | D;V | Describe the flow of participants through the study, including the number of participants with and without the outcome and, if applicable, a summary of the follow-up time. A diagram may be helpful. | Fig. 1, Tab. 1 |
| | 13b | D;V | Describe the characteristics of the participants (basic demographics, clinical features, available predictors), including the number of participants with missing data for predictors and outcome. | Fig. 1, Tab. 1 |
| | 13c | V | For validation, show a comparison with the development data of the distribution of important variables (demographics, predictors and outcome). | Tab. S5 |
| Model development | 14a | D | Specify the number of participants and outcome events in each analysis. | Tab. S5 |
| | 14b | D | If done, report the unadjusted association between each candidate predictor and outcome. | (Tab. S3/4) |
| Model specification | 15a | D | Present the full prediction model to allow predictions for individuals (i.e., all regression coefficients, and model intercept or baseline survival at a given time point). | Tab. S6, Provided Code |
| | 15b | D | Explain how to the use the prediction model. | Tab. S6, Provided Code |
| Model performance | 16 | D;V | Report performance measures (with CIs) for the prediction model. | Tab. 2,3, Tab. S8/9 |
| Model-updating | 17 | V | If done, report the results from any model updating (i.e., model specification, model performance). | NA |
| **Discussion** | | | | |
| Limitations | 18 | D;V | Discuss any limitations of the study (such as nonrepresentative sample, few events per predictor, missing data). | 14 |

| | | | | |
|---|---|---|---|---|
| Interpretation | 19a | V | For validation, discuss the results with reference to performance in the development data, and any other validation data. | 9, 10 |
| | 19b | D;V | Give an overall interpretation of the results, considering objectives, limitations, results from similar studies, and other relevant evidence. | 14 |
| Implications | 20 | D;V | Discuss the potential clinical use of the model and implications for future research. | 14, 15 |
| **Other information** | | | | |
| Supplementary information | 21 | D;V | Provide information about the availability of supplementary resources, such as study protocol, Web calculator, and data sets. | 5 |
| Funding | 22 | D;V | Give the source of funding and the role of the funders for the present study. | 14 |

**Supplementary Table 10: Tripod Statement.**

### References


1. Shah RA, Asatryan B, Sharaf Dabbagh G, Aung N, Khanji MY, Lopes LR, van Duijvenboden S, Holmes A, Muser D, Landstrom AP, Lee AM, Arora P, Semsarian C, Somers VK, Owens AT, Munroe PB, Petersen SE, Chahal CAA, Genotype-First Approach I. Frequency, Penetrance, and Variable Expressivity of Dilated Cardiomyopathy-Associated Putative Pathogenic Gene Variants in UK Biobank Participants. *Circulation* 2022;**146**:110-124.
2. Shah S, Henry A, Roselli C, Lin H, Sveinbjornsson G, Fatemifar G, Hedman AK, Wilk JB, Morley MP, Chaffin MD, Helgadottir A, Verweij N, Dehghan A, Almgren P, Andersson C, Aragam KG, Arnlov J, Backman JD, Biggs ML, Bloom HL, Brandimarto J, Brown MR, Buckbinder L, Carey DJ, Chasman DI, Chen X, Chen X, Chung J, Chutkow W, Cook JP, Delgado GE, Denaxas S, Doney AS, Dorr M, Dudley SC, Dunn ME, Engstrom G, Esko T, Felix SB, Finan C, Ford I, Ghanbari M, Ghasemi S, Giedraitis V, Giulianini F, Gottdiener JS, Gross S, Guethbjartsson DF, Gutmann R, Haggerty CM, van der Harst P, Hyde CL, Ingelsson E, Jukema JW, Kavousi M, Khaw KT, Kleber ME, Kober L, Koekemoer A, Langenberg C, Lind L, Lindgren CM, London B, Lotta LA, Lovering RC, Luan J, Magnusson P, Mahajan A, Margulies KB, Marz W, Melander O, Mordi IR, Morgan T, Morris AD, Morris AP, Morrison AC, Nagle MW, Nelson CP, Niessner A, Niiranen T, O'Donoghue ML, Owens AT, Palmer CNA, Parry HM, Perola M, Portilla-Fernandez E, Psaty BM, Regeneron Genetics C, Rice KM, Ridker PM, Romaine SPR, Rotter JI, Salo P, Salomaa V, van Setten J, Shalaby AA, Smelser DT, Smith NL, Stender S, Stott DJ, Svensson P, Tammesoo ML, Taylor KD, Teder-Laving M, Teumer A, Thorgeirsson G, Thorsteinsdottir U, Torp-Pedersen C, Trompet S, Tyl B, Uitterlinden AG, Veluchamy A, Volker U, Voors AA, Wang X, Wareham NJ, Waterworth D, Weeke PE, Weiss R, Wiggins KL, Xing H, Yerges-Armstrong LM, Yu B, Zannad F, Zhao JH, Hemingway H, Samani NJ, McMurray JJV, Yang J, Visscher PM, Newton-Cheh C, Malarstig A, Holm H, Lubitz SA, Sattar N, Holmes MV, Cappola TP, Asselbergs FW, Hingorani AD, Kuchenbaecker K, Ellinor PT, Lang CC, Stefansson K, Smith JG, Vasan RS, Swerdlow DI, Lumbers RT. Genome-wide association and Mendelian randomisation analysis provide insights into the pathogenesis of heart failure. *Nat Commun* 2020;**11**:163.
